# iTBS induces a suppression-rebound pattern in human cortical excitability

**DOI:** 10.64898/2026.09.15.26363149

**Authors:** Ethan A. Solomon, Umair Hassan, Charles W. Dickey, Christopher C. Cline, Sofia Pantis, Ritika Garg, Ashwin Ramayya, Vivek P. Buch, Matthew A. Howard, Joshua R. Tatz, Nicholas T. Trapp, Aaron D. Boes, Corey J. Keller

**Author notes:** **Correspondence:** Ethan A. Solomon, Dept. of Psychiatry and Behavioral Sciences, Stanford University School of Medicine.

## Abstract

Intermittent theta burst stimulation (iTBS) is a widely used form of brain stimulation, developed on the premise that it promotes long-term potentiation (LTP) and neural excitation. However, this is not validated in higher-order neocortex, and evidence suggests its excitatory properties may have been overstated. To address this, we recruited 20 neurosurgical patients to undergo sessions of intracranial iTBS to varied targets. We probed changes in cortical reactivity with single pulse electrical stimulation, finding that iTBS resulted in approximately 1 minute of decreased cortical reactivity in the theta and high-frequency ranges in regions strongly connected to the stimulation site. However, we observed a subsequent rise in reactivity leading up to the next iTBS session with parallel changes in resting state excitation/inhibition balance. Finally, we generalized these findings to 3 neurosurgical patients who underwent iTBS delivered by transcranial magnetic stimulation, demonstrating a similar suppression-rebound pattern. Our results suggest iTBS promotes a dynamic process of cortical suppression followed by rebound increases in excitability.

## Introduction

Brain stimulation is now an important approach in treating neuropsychiatric disorders, but in one way it is like many of the psychiatric treatments that came before it: we can use it to effectively treat brain illness even though we do not know how it works to change the brain. Transcranial magnetic stimulation (TMS) is a prime example – repeated patterns of rhythmic stimulation delivered to the dorsolateral prefrontal cortex (DLPFC) relieves symptoms of depression^1–3^, putatively through induced neuroplasticity and altered excitability of targeted brain networks^4,5^. However, the mechanism of this change is not understood, nor is the relationship between rhythmic stimulation and neuroplasticity firmly established. Elucidating this mechanism would (1) provide a window into the fundamental nature of activity-dependent neuroplasticity and (2) offer a roadmap for optimizing stimulation protocols and improving clinical benefit.

*In-vitro* physiology and animal studies first established that specific patterns of stimulation – most notably intermittent theta burst stimulation (iTBS) – caused long-term potentiation (LTP) and therefore shifted excitatory/inhibitory (E/I) balance towards excitation^6–8^. Early studies bore this out in humans by testing the effect of iTBS on motor cortex, where excitatory changes can be inferred via downstream electromyography (EMG)^9–11^. Biological models of neural circuit function can also account for an excitatory effect of iTBS^12^, bolstering confidence in this hypothesis. However, it has not been empirically validated that this hypothesis holds true in higher-order cortical areas like the DLPFC. Moreover, recent studies with more robust sham controls and larger samples have failed to replicate the large excitatory effects seen in earlier studies of iTBS to motor cortex^13^. Even if it is true that iTBS induces local increases in excitability, it is not at all clear how local plasticity translates to network-level changes. Indeed, the entire premise of functionally-guided neurostimulation in depression is that DLPFC iTBS may ultimately exert an inhibitory effect on downstream subgenual cingulate neurons^14–16^, but we lack evidence for this effect.

Excitability can be measured experimentally through several approaches. Commonly, investigators will use single-pulse stimulation events to probe cortical reactivity and analyze the neural response to infer changes in excitability from pre- to post-intervention. TMS paired with electroencephalography (EEG) provides such a probe-based measure of reactivity via the TMS-evoked potential (TEP), a brief fluctuation in EEG potentials lasting several hundred milliseconds after a single TMS pulse^17–19^. While an important tool for non-invasively measuring changes in cortical excitability, study of the TEP has revealed inconsistent patterns in the effect of iTBS on cortex^13^. Both iTBS-mediated excitation and inhibition have been reported in studies of TMS to the DLPFC as well as other cortical areas^20–22^. Moreover, the auditory and somatosensory effects of TMS make it difficult to separate neuroplastic effects from sensory habituation^23^. fMRI BOLD studies have also demonstrated that patterned TMS alters activity in a broad network of regions connected to the target site^24–27^, but shifts in E/I balance cannot be easily inferred from these signals. As such, existing non-invasive approaches are limited in what they can tell us about stimulation-related neuroplasticity.

To advance our understanding of activity-dependent neuroplasticity in higher-order human neocortex, it is crucial to (1) establish whether iTBS alters excitability in targeted tissue and/or downstream regions and (2) characterize the neuroplastic response to iTBS in terms of its magnitude, timing, and manifestation at the level of population neuronal activity. Answering these questions requires a direct measure of neural activity from human cortex undergoing iTBS. Stereo-electroencephalography (sEEG) in neurosurgical patients – wherein electrodes are implanted within brain parenchyma – provides an ideal platform for such an experiment. sEEG offers direct, *in-vivo* measurements of neural activity at high spatiotemporal resolution, enabling estimates of high-frequency activity (HFA; 75-180Hz) that is thought to correlate with multi-unit activity and changes in excitability^28–30^. Electrical stimulation can be delivered through these same electrodes, allowing us to assess the effects of iTBS targeted with high anatomical precision.

In this manuscript, we report results from 20 neurosurgical subjects who underwent sEEG recording and up to 3 sessions of direct electrical iTBS to diverse targets across the brain. To characterize the neuroplastic effect of iTBS, we analyzed the spectral perturbations provoked by single-pulse stimulation, providing a measure of cortical reactivity. Stronger responses, particularly in the HFA band, would reflect an underlying increase in cortical excitability. Next, we examined resting-state periods for evidence of changes in E/I balance independent of single-pulse probes. Finally, we generalized our findings to a cohort of 3 neurosurgical patients who received stimulation with TMS, allowing us to ask whether cortical responses to stimulation that we observed reflect a general feature of brain function versus specific responses to certain stimulation modalities.

Through this approach, we discovered that iTBS causes a pattern of suppression followed by rebound enhancement of cortical reactivity. Within seconds following a dose of iTBS, we found that single-pulse related power in the theta and HFA bands was suppressed below baseline, specifically within the network of regions strongly connected to the stimulation site. However, activity in this network rebounds within minutes, eventually increasing to surpass baseline levels. We show evidence that subsequent sessions of iTBS drive stronger suppression-to-enhancement of cortical reactivity, reflecting a potential metaplastic effect. In resting state, we quantified the exponential of the power spectrum and found iTBS-related enhancement of cortical excitability using this alternative measure. Finally, we found that these effects generalized to TMS – neurosurgical patients who underwent TMS-delivered iTBS to the DLPFC showed similar suppression-to-enhancement dynamics in the theta and HFA bands. Collectively, these data suggest that iTBS ultimately enhances excitability within the connected network, but recovery from each dose is a dynamic process involving initial periods of relative inhibition.

## Results

In this study, we delivered up to 3 sessions of direct electrical iTBS to neocortical areas in 20 neurosurgical patients, asking how iTBS affects spectral signatures of cortical reactivity and excitability. Single pulse electrical stimulation (spES) was delivered before and after each iTBS session to probe cortical reactivity (Figure 1a-b). We focused our analyses on recording channels that were “in-network” with the stimulation site, defined as those channels which exhibited high-amplitude cortico-cortical evoked potentials (CCEPs; Figure 1c-e) at baseline. Of note, non-cortical channels were also included in our analysis under the same amplitude criteria, but the terms “CCEP” and “cortical reactivity” are used throughout this manuscript for simplicity, as cortical sites comprised the bulk of responses (Supplemental Table 1). Intracranial EEG (iEEG) signal surrounding each spES event was spectrally decomposed into frequency bands of interest and power values were compared between pre-iTBS and post-iTBS events to generate a *t*-statistic measure of iTBS-related change (Figure 1f-h). Our primary analyses concentrated on HFA, a correlate of population-level neuronal spiking activity that may best reflect changes in cortical excitability^28,29^.

**Figure 1.**
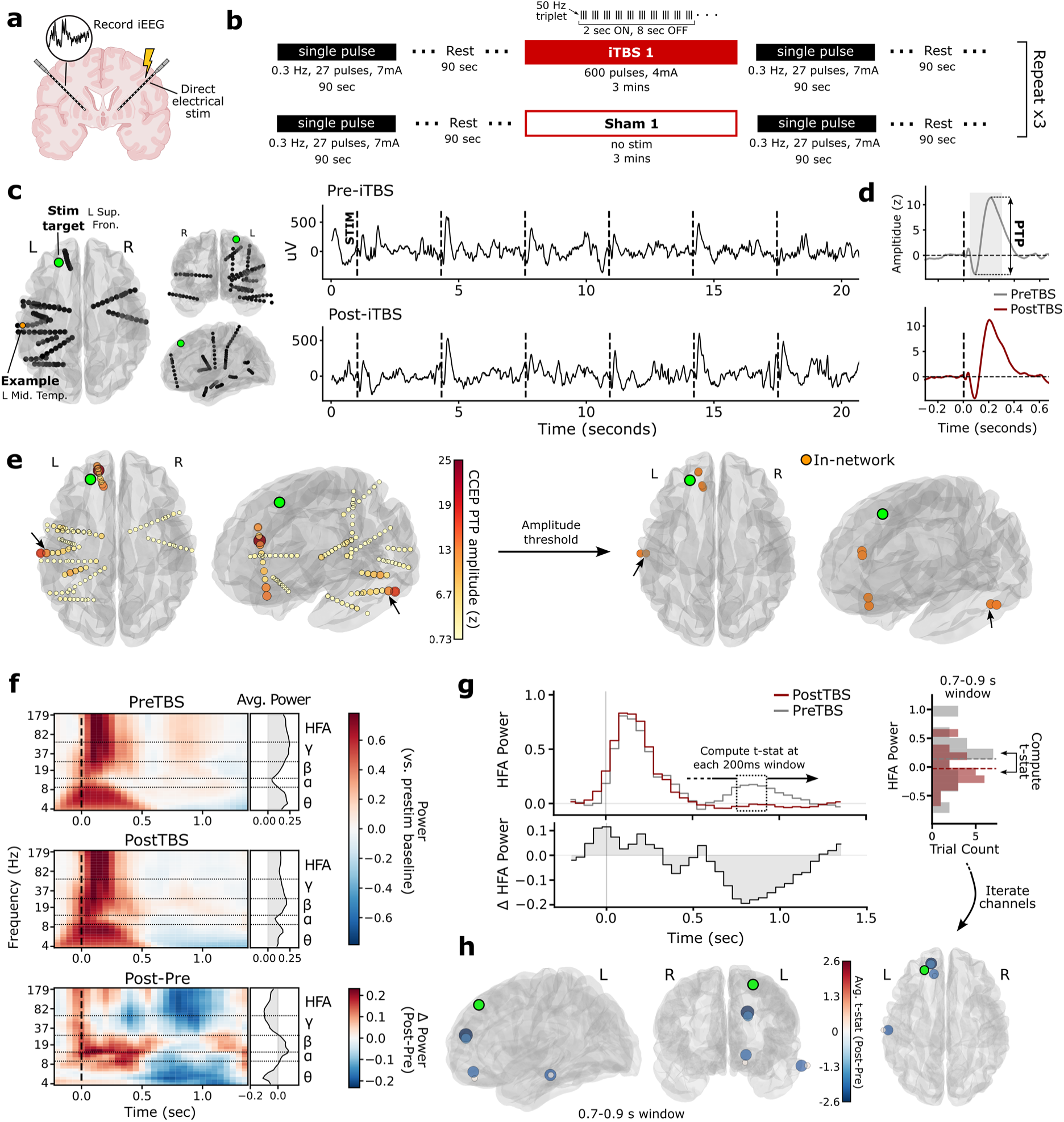
Analysis and processing pipeline in an example subject. **(a-b)** Overall task design. For a given stimulation channel, blocks of single pulses were alternated with intermittent theta burst stimulation (50Hz triplets delivered in 5Hz bursts, on for 2 seconds and off for 8 seconds, repeated 20 times) or sham (no-stimulation resting periods). For most participants, single pulses occurred in blocks of 27 pulses delivered at 7mA, at a rate of 0.3 Hz for 90 seconds. This structure was repeated 3 times, for 3 total iTBS sessions, 3 total sham sessions, and 12 total single-pulse blocks. **(c)** In an example subject, stimulation was delivered at the left superior frontal gyrus. Subsequent data in Figure 1 are derived from a recording channel in the left middle temporal gyrus. Example iEEG traces from the recording channel are shown, with single pulse stimulation events (“trials”) marked as dashed lines. The top row shows pre-iTBS events, while the bottom row shows post-iTBS events. **(d)** iEEG following each single pulse stimulation event was baseline corrected and averaged across all stimulation events in the block to generate an average waveform reflecting the cortico-cortical evoked potential, or CCEP. CCEPs were computed for the first block of single pulses given prior to any iTBS (shown here for both pre and post-iTBS for illustrative purposes). CCEP amplitude was captured as the peak-to-peak (PTP) amplitude of the evoked potential (50-300ms). **(e)** CCEP amplitude rendered for each recording channel, reflected in size and color, in the example subject. “In-network” channels are identified by setting a PTP amplitude threshold on the pre-TBS CCEPs. A threshold of 10 *z*-score units is used throughout the manuscript, while robustness to alternative thresholds is demonstrated in the Supplement. **(f)** Time-frequency representations of the spectral response to single-pulse stimulation at the example channel, averaged across all pre-TBS and post-TBS trials (top two rows). Power values were averaged into partially overlapping 200 ms windows spanning the trial, spaced 50 ms apart. Right sidecar plots show the average power at each frequency in the 0-1.5 s window. *Bottom*: Difference between post- and pre-TBS spectral power. **(g)** High-frequency activity (HFA) was computed as the average power in the 75-180 Hz band (left). For each 200ms window, the pre- and post-TBS power values are compared using a 2-sample *t*-test, generating a *t-*value for each point in time reflecting the degree to which HFA power changed from before to after the TBS session. Higher *t-*statistics indicate greater post-TBS power. The 0.7-0.9 window is shown as an example. **(h)** *T*-statistics are generated for each point in time, for each in-network electrode. Graphic in panel (a) created in BioRender.

### Cortical reactivity is initially suppressed following iTBS

After thresholding for in-network channels, 12 of the original 20 subjects had at least one in-network channel to pursue further analysis, with 71 channels among the original 2772 included (approximately 2.5%; see Figure 2a, Supplemental Table 1). Among that subset, HFA power demonstrated a sharp increase immediately following single-pulse stimulation in both the pre- and post-iTBS blocks, replicating prior work^31^. However, post-iTBS HFA was lower beginning around 400 ms (Figure 2b). The post-minus-pre (“Post-Pre”) difference in power was significant in the time windows centered on 700 ms, 750 ms, 800 ms, and 850 ms (linear mixed-effects model, *p*<0.05 after FDR correction) with the minimum achieved at 750 ms (Intercept=-0.392 [-0.625, -0.158], Wald *z*=-3.29, corrected *p*=0.022), indicating decreased cortical HFA reactivity after an intervening session of iTBS.

**Figure 2.**
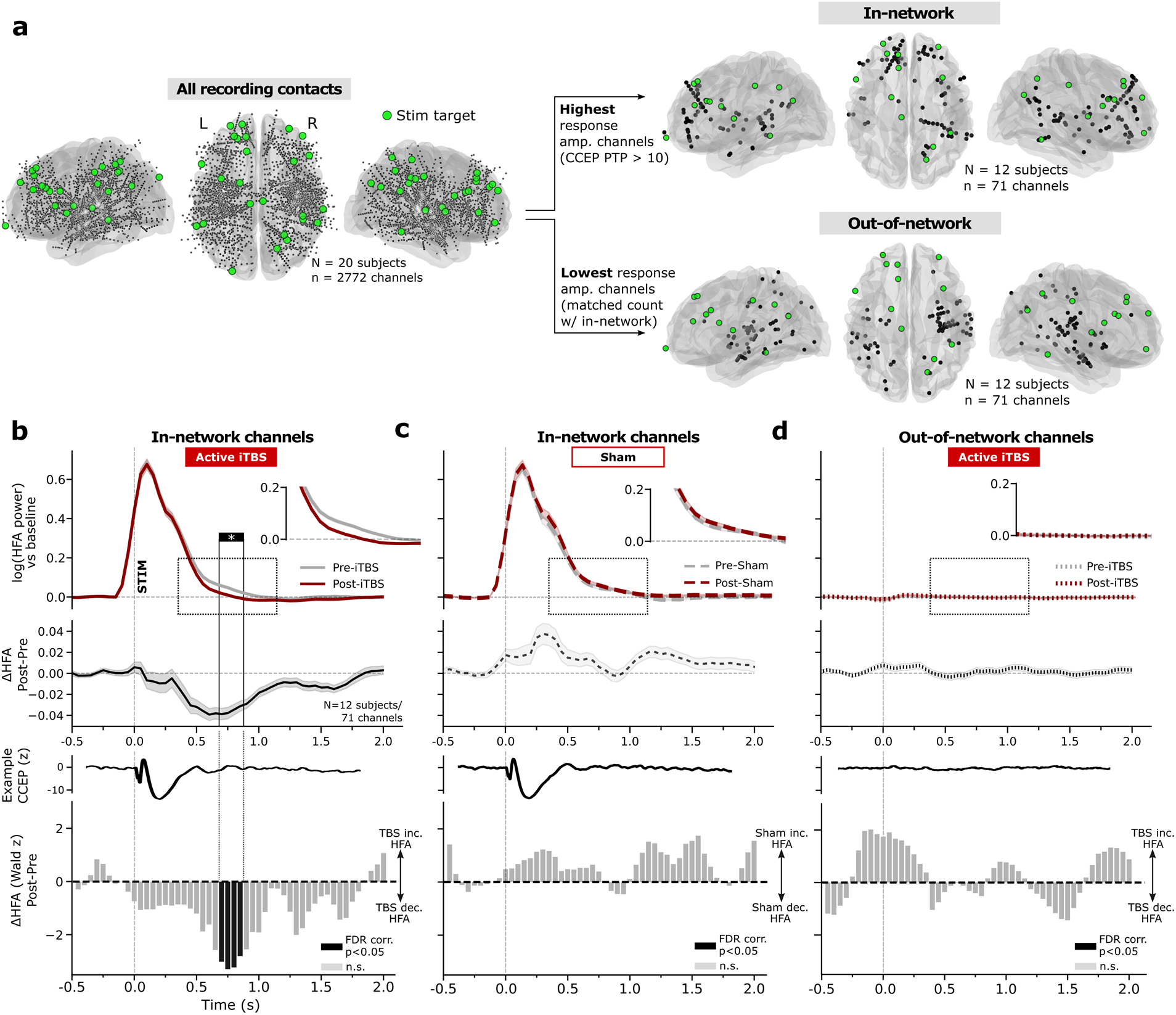
Cortical reactivity in the HFA band is reduced from pre- to post-TBS. **(a)** *Left:* All stimulation sites (green) and recording channels (black) across the 20 subjects included in this study. Stimulation sites spanned targets predominantly in prefrontal and parietal areas (see Supplemental Table 1). *Right:* In-network channels were identified as those that demonstrated high-amplitude CCEP responses to spES. A PTP threshold of 10 (*z* units) is used for figures in the main text, while alternative thresholds are demonstrated in the Supplement. Out-of-network channels were identified by selecting the channels with the lowest-amplitude CCEPs, matched in per-subject count to the number of in-network channels. **(b)** *Top to bottom:* The average timecourse of pulse-provoked HFA power in pre-(gray) and post-iTBS (red) conditions was computed across the 71 channels in 12 subjects that were in-network, combining effects across all iTBS sessions. Power appears to rise prior to stimulation due to temporal smoothing from spectral windowing and temporal binning described in *Methods*; no significant effects were observed in any windows overlapping the stimulation pulse. Post-Pre difference in HFA is shown for each timepoint, averaged across all channels and participants. Error bars show +/-1 SEM across all pooled channels. An example CCEP from a single channel among the in-network group is shown to demonstrate correspondences between HFA effects and canonical CCEP components. *Bottom:* Given the variable number of contacts included per subject, a linear mixed-effects modeling approach was used to estimate the population-level effect at each timepoint, generating a Wald *z* statistic (see *Methods* for details). After FDR correction for multiple comparisons, a significant Post-Pre decrease in HFA was observed at 0.7 s, 0.75 s, 0.8 s, and 0.85 s. **(c)** As in (b), demonstrating post-sham minus pre-sham effects among in-network channels, averaged across all sham sessions. No significant sham-related change in HFA power was noted. **(d)** As in (b), demonstrating Post-Pre iTBS effects for the group of out-of-network channels (low effective connectivity to the stimulation site). No significant change in HFA power was noted in this group.

As a control, we looked for HFA changes between pre- and post-sham single pulse blocks, within the same set of in-network channels. As active intracranial iTBS is itself imperceptible, sham periods were simply 3-minute periods of rest with no stimulation events. No significant change in HFA power was observed at any time bin when comparing pre- and post-sham trials (Figure 2c; maximum at 1.55 s; Intercept=0.154 [-0.019, 0.328], Wald *z*=1.746, corrected *p*=0.86). The trend towards sham-related increases in HFA is explored in more depth later in this manuscript. The Post-Pre effect of iTBS on HFA was significantly different between active and sham in the 0.7-0.85 s time window identified above (Wald *z*=-5.50, *p*<0.001). We also evaluated HFA changes in “out-of-network” channels, which is the group of channels showing the lowest CCEP responses and matched in per-subject count to the in-network electrodes (see *Methods*, Figure 2a). No significant change from pre-iTBS to post-iTBS was observed in any time bin in the out-of-network group of channels (Figure 2d; maximum at 50ms; Intercept=0.202 [-0.005, 0.410], Wald *z*=1.908, corrected *p*=0.83) and there was a significant difference between in-network and out-of-network effects in the 0.7-0.85 s window (*z*=-2.41, *p*=0.016).

This finding of iTBS-related suppression of HFA power was stable across thresholds used to determine the set of in-network channels, including substantially lower cutoff values (Supplemental Figure 1). Significant iTBS-related decrease in HFA was observed in the 0.7-0.85 s window starting at a CCEP amplitude threshold of 5, which includes a total of 613 recording channels (Figure 3a and Supplemental Figure 2a-b; approximately 22% of total) across 15 subjects and 17 unique stimulation sites (counting two subjects with two targets each).

**Figure 3.**
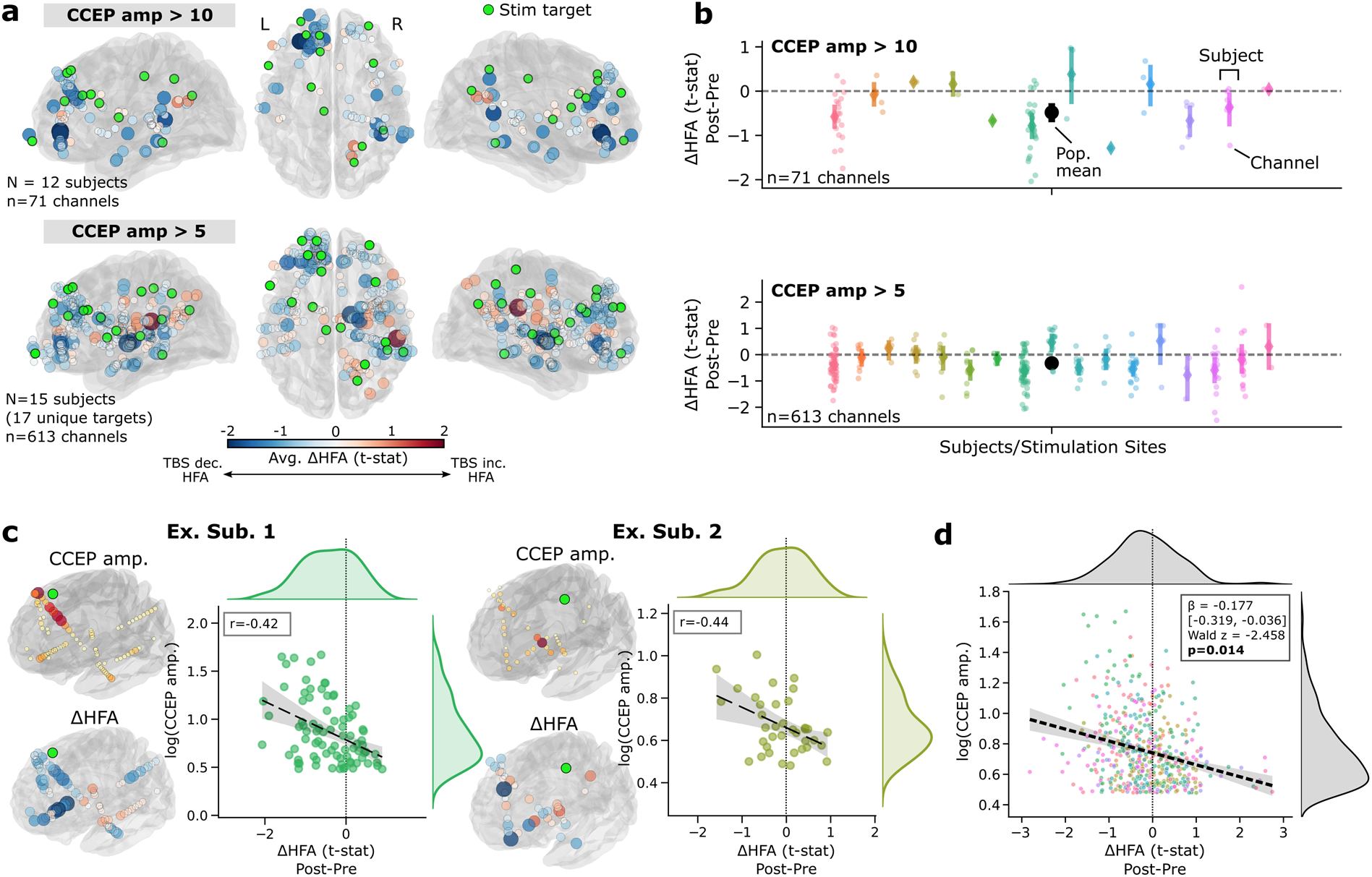
Change in cortical reactivity is inversely correlated with CCEP amplitude. **(a)** *Top:* The 71 recording channels across 12 participants that met CCEP amplitude threshold >10 (“in-network” channels), colored by the Post-Pre HFA t-statistic in the 0.7-0.85 s time window identified previously. Blue colors reflect greater HFA decreases from pre to post iTBS. *Bottom*: The 613 recording channels included as “in-network” under a >5 threshold, colored same as above. **(b)** Rain plots depicting each individual recording contact as a point, colored by subject. Negative values indicate Post-Pre iTBS decreases in HFA power, in the 0.7 to 0.85 s window. Distributions are shown for CCEP amplitude thresholds at 10 (*top*) and 5 (*bottom*; see Supplemental Figures for further analysis). Pooled population-level mean and SEM are indicated in black at the center of each plot. **(c)** Correlation between Post-Pre change in HFA power (ΔHFA) in the 0.7-0.85 s window and CCEP amplitude, for each contact within 2 example subjects chosen to demonstrate the effect. Data reflects all contacts with a CCEP amplitude greater than 3 (z-score units), to better capture variability across a wide range of responses. See Supplemental Figure 3 for all included subjects. **(d)** Same as (C), aggregated across all included participants (N=17 with at least 5 channels included under the >3 threshold). Across the population, there is a significant (*p*=0.014) inverse relationship between ΔHFA and CCEP amplitude, as estimated by a mixed-effects model that accounts for Euclidean distance.

Included stimulation sites spanned a range of brain regions, though the most common stimulation areas used were prefrontal and parietal cortices from either hemisphere (Figure 3a). In-network channels spanned diverse regions including prefrontal/cingulate, temporal, parietal, and thalamic sites (see Supplemental Table 1 for further detail). While most in-network channels demonstrated an iTBS-related HFA decrease in the 0.7-0.85 s window, we observed a range of response magnitudes at both low and high CCEP amplitude cutoffs (Figure 3b). We hypothesized that the degree of HFA suppression observed for each channel could be related to the strength of its connectivity to the stimulation site. For this specific test, we adopted a lower amplitude threshold of 3 to best capture variability across high and low responses.

Both within individual subjects and across the population, HFA suppression at a given channel was negatively correlated with CCEP amplitude – larger CCEPs (and therefore stronger effective connectivity) yielded greater HFA suppression (Figure 3c-d; population analysis: β=-0.177, Wald *z*=-2.458, *p*=0.014 by mixed effects model, see Supplemental Figure 3 for individual subjects and *Methods* for details). Our model accounts for Euclidean distance between stimulation sites and response channels, which is itself marginally negatively correlated with HFA suppression (β=-0.152, Wald *z*=-1.945, *p*=0.052); therefore, CCEP amplitude predicts HFA response beyond what would be expected by distance to the stimulation site alone. We also note that this inverse relationship is observed at multiple channel inclusion thresholds and reaches significance at thresholds of 3 and 5 (Supplemental Figure 4). Taken together, this analysis indicates that channels with stronger connectivity to the stimulation site exhibited greater iTBS-related reduction in cortical reactivity.

### iTBS predominantly modulates theta and HFA

While HFA may best correlate with cortical excitability, prior work suggests iTBS may also affect oscillatory activity at lower frequencies^32–34^. We therefore asked whether iTBS-related reduction in cortical reactivity was limited only to HFA or generalized to other frequency bands. In the population-level average both before and after iTBS, spES provoked a strong broadband spectral response within the first 500 ms after stimulation (Figure 4a). However, in the 0.7-0.85 s interval identified previously – which occurs much later than the large-amplitude initial response – spectral power effects are inverted from pre to post-iTBS; theta (4-8 Hz), alpha (9-13 Hz), beta (15-25 Hz), and HFA are elevated above baseline in pre-iTBS trials but fall below baseline in post-iTBS trials. This differential is significant in theta, alpha, and HFA (*p*<0.05 by mixed-effects model, Bonferroni corrected). The gamma (30-55 Hz) and beta (15-25 Hz) bands showed nonsignificant change from pre to post-stimulation. Given that the alpha and theta effects are similar and may reflect spectral overlap between these bands, we focus subsequent analyses in this manuscript on theta and HFA only.

**Figure 4.**
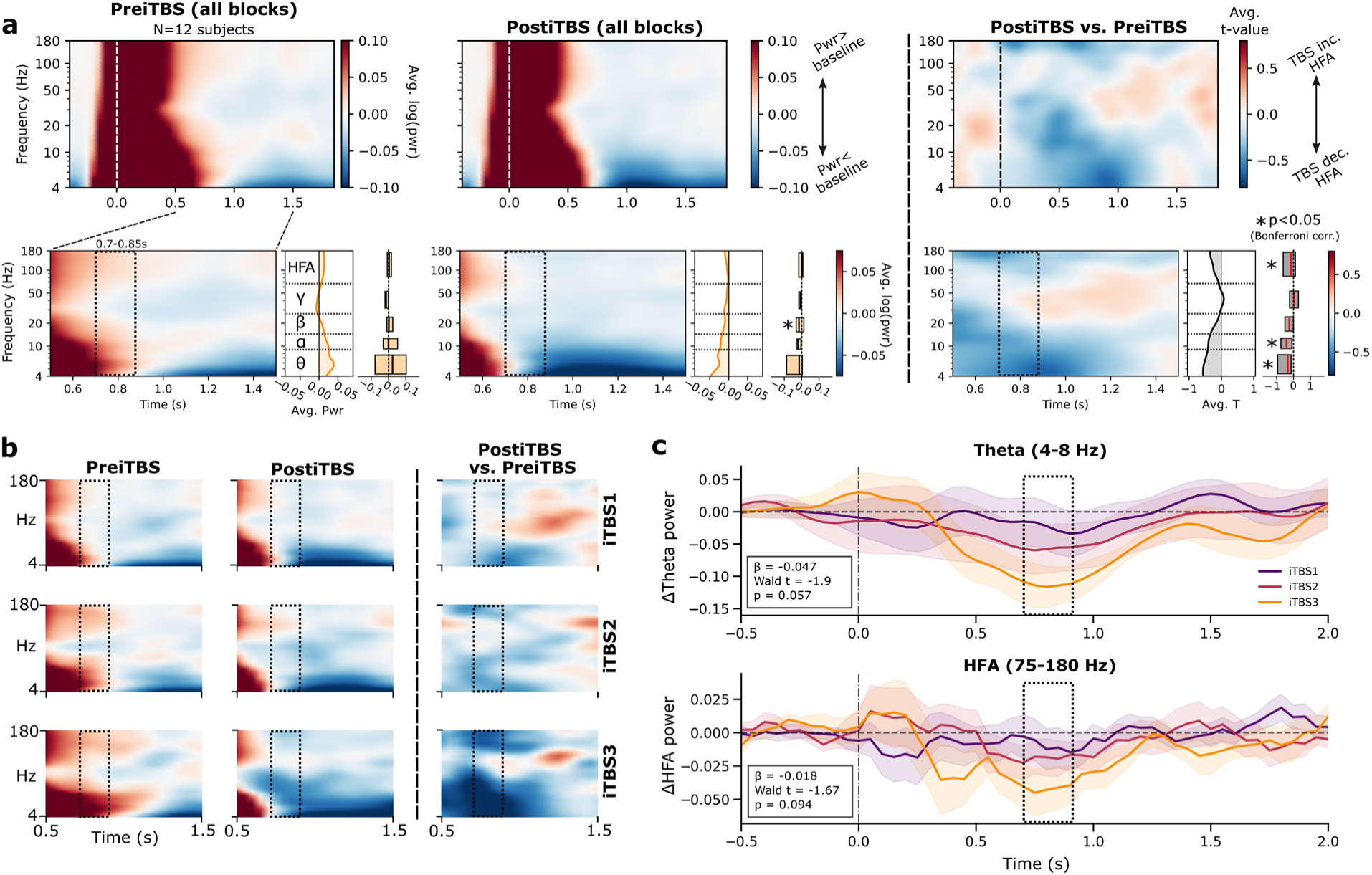
iTBS predominantly modulates cortical reactivity in the theta and HFA bands. **(a)** Grand-average TFRs showing the spectral response following single-pulse stimulation, averaged across all pre-iTBS or post-iTBS trials (for all iTBS sessions), and then averaged across all in-network recording channels for all subjects. Blue colors reflect power decreases relative to the pre-pulse baseline. Bottom row shows a focus window from 0.5 s to 1.5 s, with the 0.7-0.85 s interval of significant iTBS modulation (see Figure 2) highlighted. Sidecar plots show the average spectral power within the 0.7-0.85 s window, in key frequency bands of interest. Boxes indicate the interquartile range across subjects. Rightmost TFR shows the average t-statistic reflecting the Post-Pre change in spectral power. Blue colors reflect decreases in spectral power from before to after iTBS. \**p*<0.05 (Bonferroni corrected) difference from zero, estimated by a linear-mixed effects model (see *Methods* for details). **(b)** TFRs shown for each iTBS session (1-3) individually, otherwise structured as in (A). **(c)** Band-averaged post vs. pre-iTBS power in the HFA and theta (4-8 Hz) ranges plotted over time, demonstrating a progression of effect size through each iTBS session. Box indicates the 0.7-0.85 modulation window of interest. In this window, a test for linear progression of the effect size through blocks showed a trend (p<0.1) for both theta and HFA (see inset boxes for statistics).

As our protocol implemented three sessions of iTBS with interleaved single-pulse probes, we next asked whether changes in cortical reactivity progressed with successive iTBS doses. In both theta and HFA, there was a progression of power decreases with each iTBS session, such that the degree of power suppression was least following iTBS1 and greatest following iTBS3 (Figure 4b-c). For both theta and HFA, this suppression effect met significance at the third iTBS session (HFA: Wald *z*=-3.074, *p*=0.002; theta: Wald *z*=-3.954, *p*<0.001; Figure 5a). When testing for a linear relationship between iTBS block and power we found a correlation that was subthreshold for significance (theta: β=-0.047, Wald *z*=-1.9, *p*=0.057; HFA: β=-0.018, Wald *z*=-1.67, *p*=0.094; Figure 4c). See Supplemental Figure 5 for TFRs reflecting sham-related change in cortical reactivity.

**Figure 5.**
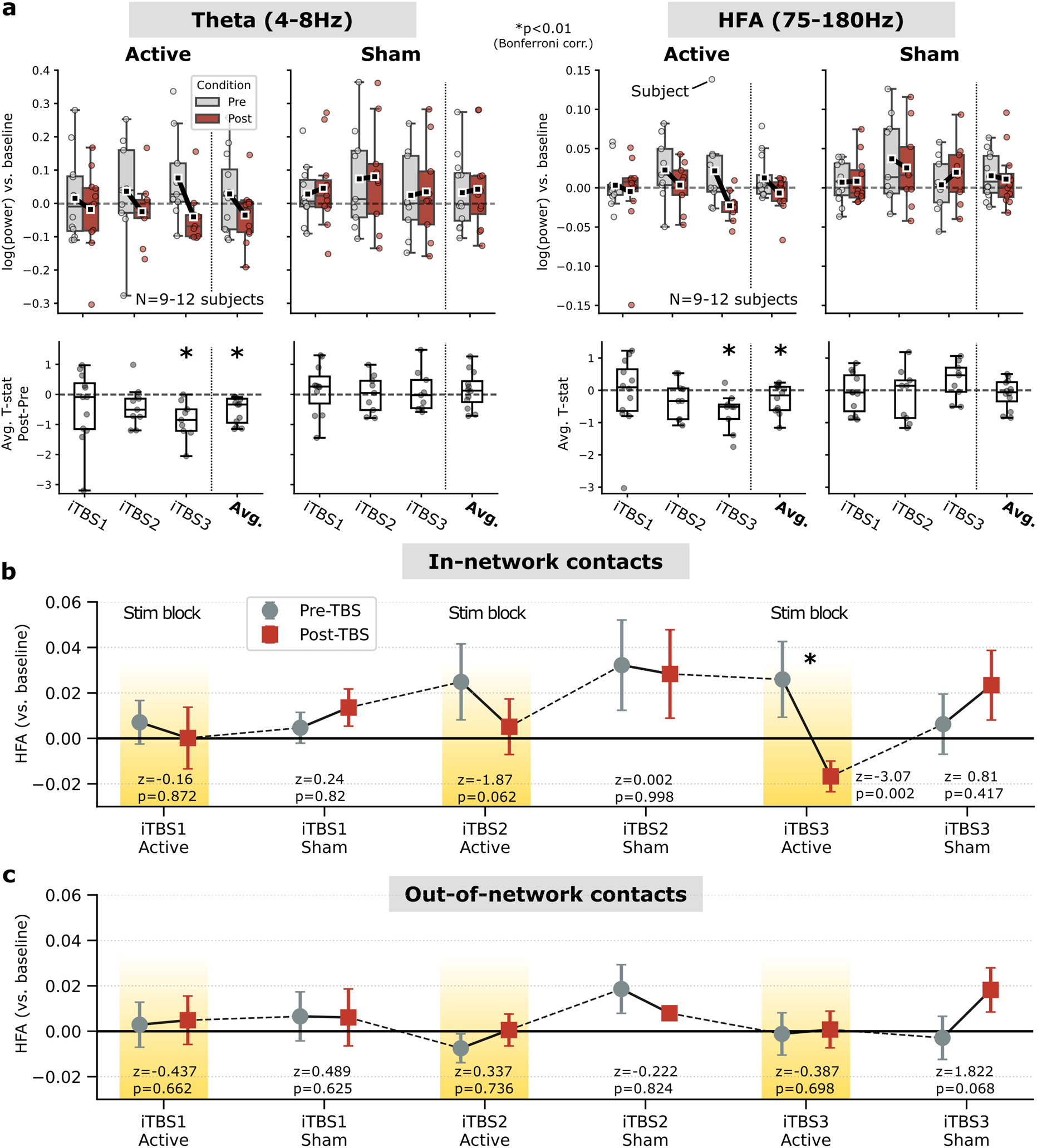
HFA power evolves across multiple iTBS sessions. **(a)** *Top:* Pre- and post-iTBS power – provoked by CCEPs – was individually measured for each block of the entire experimental session, spanning 3 active iTBS blocks and 3 “sham” periods in which no stimulation was delivered. Given the population-level effect observed in the 0.7-0.85 s window, we focused this analysis on that same period. For active iTBS conditions, HFA and theta power decreased with each iTBS session, while no significant change was observed from pre- to post-sham sessions. However, spectral power was elevated above baseline for both pre-sham and post-sham CCEP blocks. *Bottom*: Average post vs. pre *t*-statistics for each subject, broken down by iTBS session. In both theta and HFA bands, there was a significant population-level decrease in power observed by iTBS session 3 as well as in the grand average across all sessions (*p*<0.01). No significant effects were observed for sham stimulation sessions. 12 subjects contributed data to iTBS1; 9 subjects each contributed to iTBS2-3 due to variability in experimental procedures; see *Methods* and Supplemental Table 2 for details. **(b)** *Top:* Data from (A) arranged in chronological sequence, providing an overview of changes in HFA power over the course of an entire experiment. iTBS sessions resulted in initial HFA power decreases, followed by increases above baseline as time passed. Note that error bars show +/- 1 SEM across subjects, but statistical tests do not map directly to this measure of variance due to the hierarchical nature of the mixed-effects test. **(c)** Same as (b), for the set of 71 channels across the same 12 subjects that exhibited the weakest or absent CCEP response (“out-of-network”). No significant iTBS-related modulation was observed at out-of-network contacts for any session or in the grand average.

### Cortical excitability rebounds in the minutes following iTBS sessions

Collectively, our results so far point to an unexpected phenomenon: iTBS appears to decrease cortical reactivity to stimulation, at least as estimated via spES provoked theta and HFA power. Moreover, this effect correlates with the strength of connectivity to the stimulation target, and may progress through iterative sessions of iTBS. No such suppression effect was observed when we examined pre/post sham periods, in which subjects are not undergoing any form of stimulation for 3 minutes (active stimulation is imperceptible and therefore not matched with any other kind of stimulus during sham periods). Curiously, however, we observed that HFA increased from pre to post-sham at most post-pulse latencies (Figure 2c) – meaning that cortical reactivity may have increased over the sham interval.

To investigate this more closely, we directly examined pulse-provoked spectral power occurring in the CCEP blocks before and after sham intervals. In both bands, provoked power was elevated above baseline in the pre- and post-sham CCEP blocks (Figure 5a), even while there was no significant change between them. When visualized in temporal sequence of the experiment, we were surprised to find that HFA appeared to rebound from a state of post-iTBS suppression to elevation (Figure 5b). Specifically, we noted that HFA measured during pre-sham and post-sham blocks were universally higher than the post-active iTBS block which came before them. As a control, we looked for this phenomenon in the out-of-network group of channels (see Figure 2a). In this subset, no iTBS sessions or Sham sessions yielded any significant pre/post change in either HFA or theta (Figure 5c), nor did we note a clear rebound phenomenon. These observed dynamics are similar under the more inclusive channel threshold of *z* > 5 (Supplemental Figure 2c).

Taken together, these analyses demonstrated that cortical reactivity appeared to increase with more elapsed time following each iTBS session, from a state of relative suppression to relative activation. However, the effect was only noted in the subset of in-network channels; out-of-network channels showed no significant effect, consistent with the conclusion this was not a generalized or brain-wide phenomenon.

To quantify this post-iTBS rebound in cortical reactivity, we directly contrasted spectral power during the two blocks of CCEPs spaced by the greatest amount of time, but with no intervening iTBS session (“Post-Active” blocks versus “Post-Sham” block, Figure 6a). This provided an approximately 15-minute window of time during which we could track the “recovery” of brain networks from a session of iTBS. Such intervals occur three times throughout the experiment in most subjects. We found that HFA and theta power in the 0.7-0.85 s window increased during these recovery periods, starting at or below the pre-pulse baseline and rising above baseline by the end (Figure 6b). In the HFA band, this effect was significant following iTBS3 (Intercept=0.048 [0.018, 0.078], Wald *z*=3.132, *p*=0.002) and when pooling effects across all three blocks (Intercept=0.026 [0.005, 0.048], Wald *z*=2.451, *p*=0.014). The magnitude of this rebound effect trended upwards with successive iTBS sessions, but did not reach significance for a linear effect across blocks (β=0.015 [-0.004, 0.033], Wald *z*=1.572, *p*=0.116). No significant effect was observed when testing for a Post-Active to Post-Sham effect in the group of out-of-network channels (Intercept=0.009 [-0.002, 0.021], *z*=1.559, *p*=0.119), though the direction of change was also positive.

**Figure 6.**
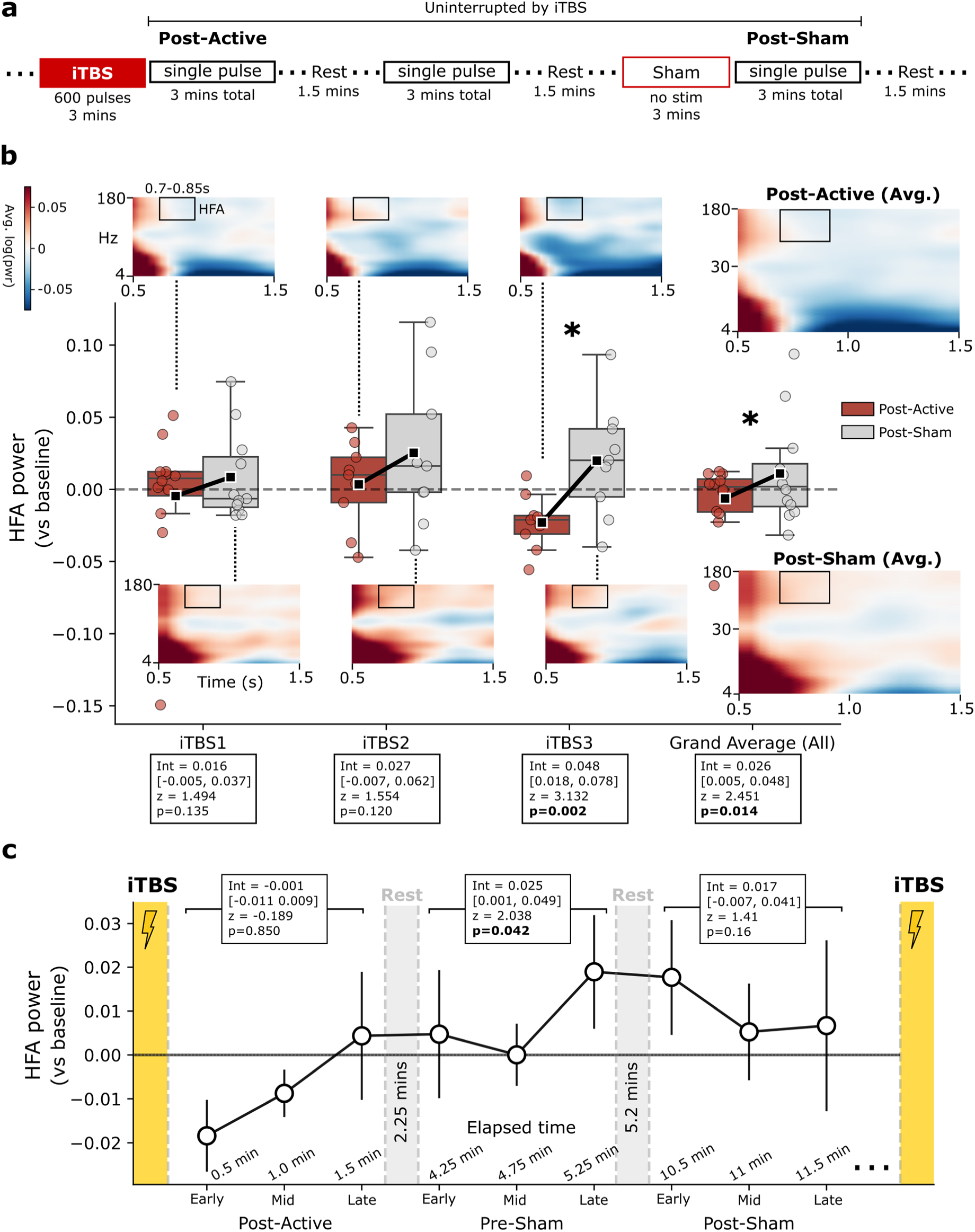
HFA increases during recovery from iTBS. **(a)** Experimental protocol schematic annotating the approximately 15-minute interval spanning the first post-iTBS block of single pulses (“Post-Active”) and the last block prior to the next iTBS session (“Post-Sham”). This interval is the longest period of time in the experiment uninterrupted by any iTBS. Note that single-pulse blocks are 3 minutes long due to a series of low-amplitude pulses also delivered for 90 s at 0.3 Hz, which were otherwise excluded from analysis in this manuscript. **(b)** In-network HFA power measured in the Post-Active (red) and Post-Sham (gray) blocks, across the 12 subjects included in prior analyses with accompanying subject-average TFRs (9 subjects each contribute to iTBS2-3). Power is shown for the 0.7-0.85 s window of interest. For each iTBS block, there is an increase in HFA power during the ∼15 minutes from the Post-Active to Post-Sham period. Averaged across all blocks, this increase is significant at *p*=0.014. **(c)** To inspect this change in HFA with greater temporal specificity, each single-pulse block (27 trials total) was divided into three sub-blocks of 9 trials each, and the average in-network HFA power was measured for each. Across the three blocks of single pulses spanning the time period between iTBS, HFA tended to increase from below baseline (Post-Active block) to above baseline (Pre-Sham and Post-Sham). Inset statistics test for 0.7-0.85 s power relative to baseline, averaged within the block; the Pre-Sham block is significantly elevated above baseline at *p*=0.042. Time indicators reflect actual time elapsed during the experiment, averaged across subjects; actual timings are shorter than per-protocol timings, often due to slight abridgment of resting intervals due to time constraints. Note that this analysis contains N=8 subjects who completed the full experimental protocol through three iTBS sessions with limited deviation from prescribed timings. Error bars show +/- 1 SEM across subjects.

In the theta band, iTBS1 and iTBS2 demonstrated significant power increases from Post-Active to Post-Sham (Wald *z*=3.729, *p*<0.001; *z*=2.991, *p*=0.003 respectively) as well as a significant pooled effect across all blocks (*z*=3.408, *p*<0.001). In both bands, pooled Post-Active to Post-Sham increases are similar when using the more liberal CCEP amplitude threshold of 5 (theta: *z*=3.10, *p*=0.002; HFA: *z*=1.67, *p*=0.095; see Supplemental Figure 2d).

To provide a higher-resolution view of this effect – asking if more prominent changes occur earlier or later in this interval – we binned each CCEP block into subgroups of 9 trials each, providing a within-block measure of HFA power (Figure 6c). We also measured experimental time while accounting for small deviations from protocol timing, to more accurately assess how power evolves between iTBS sessions. HFA was only suppressed below baseline levels in the early and middle portions of the first iTBS block (spanning about 1 minute after iTBS). Beyond this point, HFA rose above baseline, significantly so in the Pre-Sham block starting about 4 minutes after iTBS (Intercept=0.025 [0.001, 0.049], Wald *z*=2.038, *p*=0.042). Over the next 6 to 7 minutes, HFA remained elevated above baseline without reaching statistical significance (Intercept=0.017 [-0.007, 0.041], Wald *z*=1.41, *p*=0.16).

### iTBS alters resting-state measures of cortical excitability

Until this point, we have framed our findings in terms of cortical reactivity – our observation of immediate iTBS-related HFA decreases, followed by a minutes-long rebound, was captured via single-pulse probes. While their time-locked nature make single pulses a useful paradigm for exploring changes in excitability, it is possible that provoked dynamics do not represent neural dynamics without exogenous perturbations. As such, we next asked whether iTBS affected resting-state measures of cortical excitability. To do this, we estimated the exponent of the resting-state power spectrum in successive 3-second windows across the entire experimental session, during periods of time without iTBS or CCEPs (Figure 7a). The power spectrum exponent is thought to reflect excitatory/inhibitory balance, with steeper slopes (higher exponents) indicating greater inhibition, and shallower slopes (lower exponent) indicating greater excitation^35,36^. Given these properties, we considered it an ideal tool to quantify cortical excitability in the absence of any time-locked neural responses.

**Figure 7.**
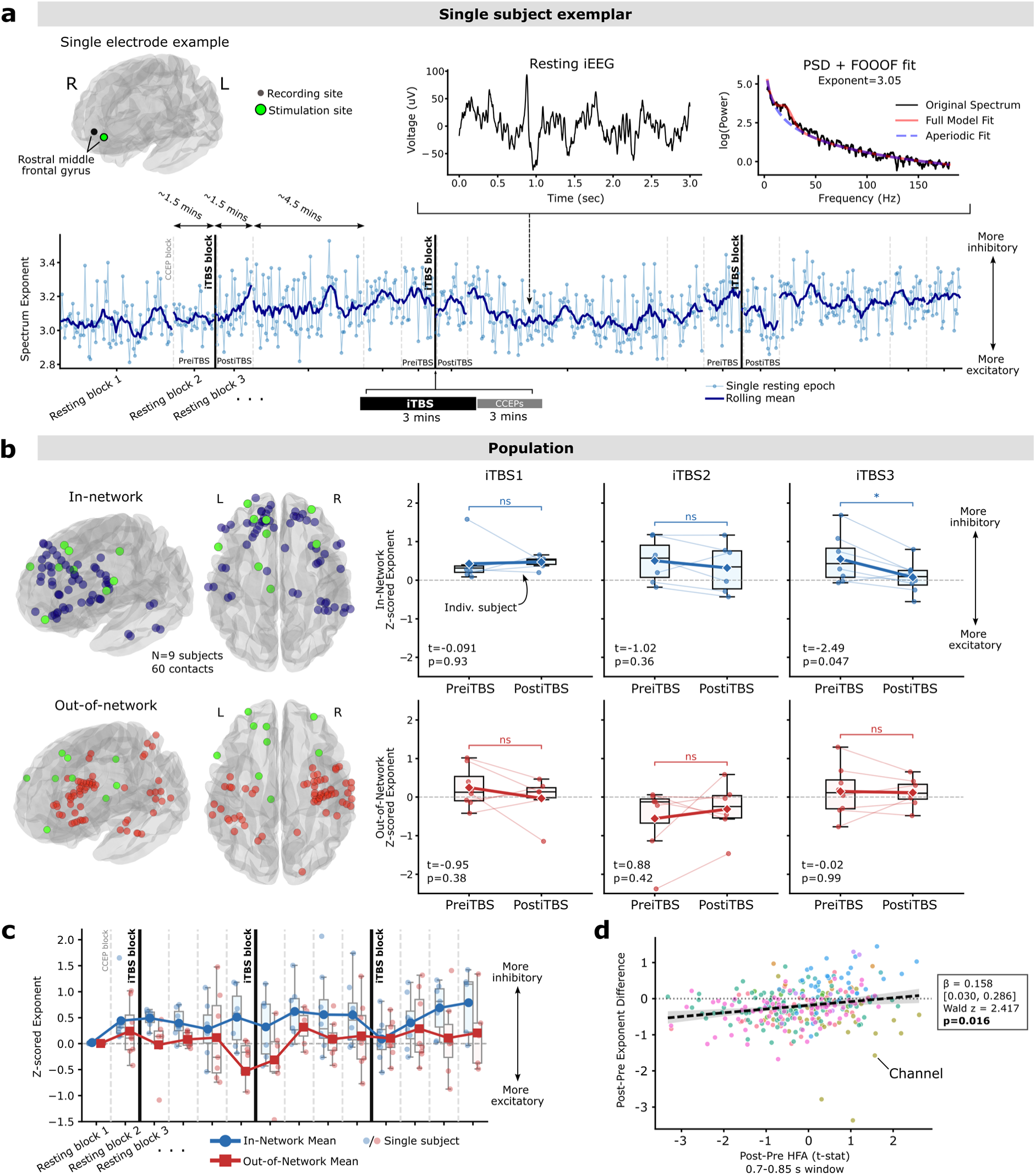
iTBS drives resting-state measures of excitation. **(a)** Resting-state analysis for an example in-network channel. For one left prefrontal electrode in an example subject, all resting-state blocks of the task (including 1.5 minute blocks between CCEP blocks and 3 minute blocks otherwise called “sham”) were epoched into successive 3-second windows. For each window, FOOOF^36^ was used to estimate the exponent of the aperiodic spectrum (upper right; see *Methods* for details). For an example electrode, the exponent estimate, and a rolling average, is plotted over all epochs and resting-state blocks in the task, with iTBS sessions marked in bold lines and CCEP sessions marked in dashed gray lines. “PreiTBS” blocks are defined as rest periods immediately prior to iTBS, while “PostiTBS” are defined as resting blocks immediately following iTBS. **(b)** Generalizing this analysis across all in-network electrodes (blue) in all participants who completed a stimulation protocol with substantial resting-state time (N=9), there was a significant main effect of TBS across all sessions (Wald *z*=-2.193, *p*=0.028) corresponding to a decrease in the aperiodic exponent. Specifically, a significant decrease in the exponent from pre-to post-iTBS is observed by the third TBS session (paired *t*-test, p=0.047, *t*(6)=-2.49). Out-of-network channels (red) do not show a significant exponent decrease for any iTBS session, and there is a significant interaction between iTBS effect and in-vs. out-of-network (LME; Wald *z*=3.064, *p*=0.002). **(c)** Average exponent (z-scored to first resting block baseline) across all resting-state blocks. In-network channels demonstrate iTBS-related changes in excitability – superimposed on a background drift towards higher exponents – while out-of-network channels show no clear trend over time. **(d)** At the iTBS3 block where significant Post-Pre in-network exponent change was detected, exponent change was positively correlated with Post-Pre HFA change derived from the earlier cortical reactivity experiment (β=0.158, *p*=0.016 by mixed effects model). These data reflect all channels with a CCEP amplitude threshold >3, to capture variability across high and low responses. Colors delineate individual subjects.

Using this approach, we asked how the spectrum exponent changes from pre to post-iTBS, examining the resting-state periods that occurred closest in time to each iTBS session (see Figure 1a and Figure 7a). Post-iTBS resting periods always occurred after the 3-minute block of post-iTBS single pulses, suggesting our earliest resting-state measure of excitability would capture the rebound response noted in Figure 6. Of note, 9 subjects were included in this analysis who completed a version of this experiment with substantial longitudinal resting-state time interleaved with iTBS blocks (see *Methods* for details).

Among the subset of in-network channels identified previously, we observed a significant decrease in the spectrum exponent from pre to post-iTBS when pooling effects across all three sessions (Intercept=-0.290 [-0.549, -0.031], Wald *z*=-2.193, *p*=0.028). The effect increased in magnitude for each subsequent iTBS session (Figure 7b), and reached significance by the third iTBS session (paired *t*-test, *t*(6)=-2.49, *p*=0.047), though this would not survive Bonferroni correction for multiple comparisons. Out-of-network channels, which exhibited little or no observable CCEP, showed no significant effect for any iTBS block (*p*>0.05) or averaged across blocks (Intercept=-0.074 [-0.349, 0.201], Wald *z*=-0.528, *p*=0.597). There was a significant difference between exponent change observed in-network versus out-of-network channels (β=0.267 [0.096, 0.438], Wald *z*=3.064, *p*=0.002). Visualized across the entire experimental session, we noted decreased exponents (signaling increased excitability) following iTBS, followed by a gradual drift back to higher exponents across subsequent resting-state blocks. The average exponent across out-of-network channels remained at baseline across the session (Figure 7c).

Finally, we observed that – at the iTBS3 block which exhibited significant exponent change – channel-wise exponent change and pulse-provoked HFA power change were significantly correlated (β=0.158 [0.030, 0.286], Wald *z*=2.417, *p*=0.016), suggesting that channels which showed greater suppression of cortical reactivity also saw greater post-iTBS resting state excitation (Figure 7d). CCEP amplitude was non-significantly negatively correlated with exponent decrease from pre to post iTBS3 (β=-0.401 [-0.848, 0.046], Wald *z*=-1.759, *p*=0.079), indicating a possibility that channels more strongly connected to the stimulation site showed greater increases in post-iTBS resting excitability (Supplemental Figure 6).

### Suppression-rebound following direct electrical iTBS generalizes to TMS-iTBS

In the preceding experiments, we used direct electrical stimulation via indwelling electrodes in the brain to show that iTBS results in an initial suppression of cortical reactivity – as measured via theta and HFA spectral activity – followed by a rebound to higher levels over several minutes. However, direct electrical iTBS is not widely used in clinical practice today. TMS is instead the mainstay of most neuromodulatory interventions in psychiatry, and iTBS delivered by TMS is FDA cleared for treatment of major depression. TMS differs substantially from direct electrical stimulation: the volume of engaged cortex is far higher^37,38^, properties of the induced electric field in the brain are sensitive to coil placement and orientation^19^, and TMS results in off-target auditory and somatosensory effects^39^. Owing to these differences, it is not clear that iTBS-related plasticity effects generalize to TMS.

To address this limitation, we recruited 3 additional neurosurgical subjects to undergo TMS-delivered iTBS to the DLPFC, while simultaneously recording from indwelling electrodes in a manner similar to what we have shown earlier in this manuscript and in prior studies^38,40–43^. Our purpose was to investigate whether the suppression-rebound dynamic we discovered in association with direct electrical stimulation was also present with TMS. Procedures to identify in-network electrodes, spectral analysis, and statistical methods were otherwise similar, although in this experiment each subject received only 1 session of iTBS, preceded and followed by blocks of 50 single pulses delivered to the same DLPFC site (see *Methods* for stimulation parameters). Additionally, we adopted a lower *z-*score threshold for in-network inclusion due to an overall lower amplitude of TMS-evoked responses (*z-*score of 3; Figure 8a-b), termed intracranial TMS-evoked potentials (iTEP). We continued to use the 0.7-0.85 s modulation window as our primary timepoint of interest, providing a strict measure of generalization.

**Figure 8.**
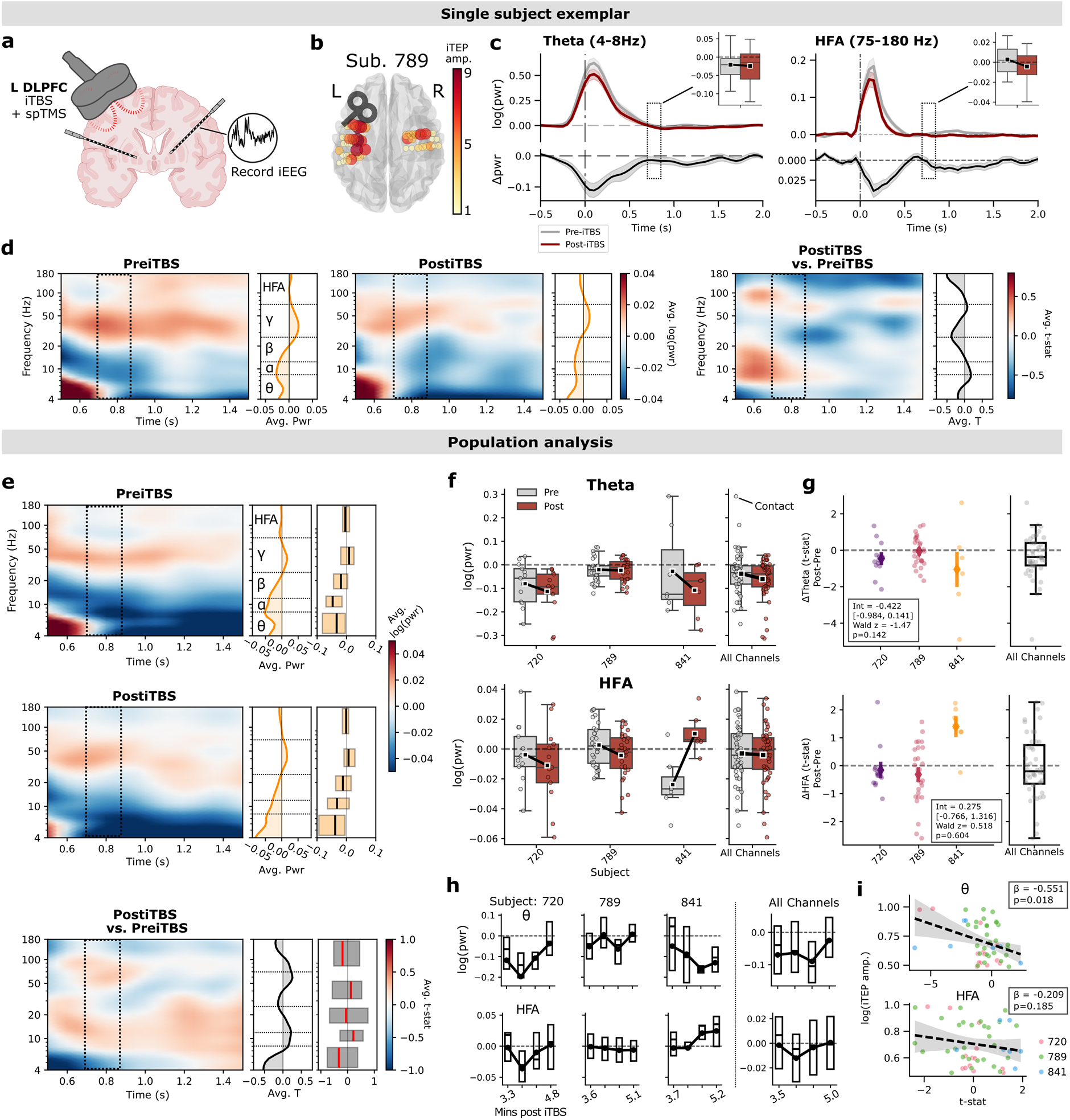
iTBS-related change in cortical reactivity generalizes to TMS. **(a)** In 3 additional participants, iTBS and single pulses were delivered via TMS to the dorsolateral prefrontal cortex (see *Methods* for details). 50 single pulses at a fixed stimulation intensity were delivered before and after a single session of iTBS. Analysis procedures were similar to those described in Figure 1 and *Methods*, aside from a lower TEP amplitude threshold necessary due to generally lower-amplitude responses with TMS. **(b)** iTEP amplitudes for all recording contacts in an example subject, reflected in the size and color of each channel marker, similar to Figure 1e. See Supplemental Figure 7 for all subjects. **(c)** spTMS-provoked theta and HFA timecourses (top) with associated post vs. pre-iTBS differences (bottom) averaged across the 28 channels that exceeded iTEP amplitude threshold (in-network), for the same example subject as in (B). Boxplots represent the distribution of power values across in-network channels, for the 0.7-0.85 s window of interest used throughout this manuscript. As seen in electrical iTBS, power decreases were observed in both theta and HFA. **(d)** Time-frequency responses to spTMS averaged across the 28 in-network channels in the same example subject, structured similarly to Figure 4a. Sidecar plots show averaged power values or Post-Pre *t*-statistics in the 0.7-0.85 s window. **(e)** Averaged time-frequency responses to spTMS across all in-network channels in the group of 3 TMS-iEEG participants. As seen in prior analyses, iTBS tended to decrease pulse-provoked HFA and theta power. **(f)** Theta (top) and HFA (bottom) power values averaged across all pre and post-iTBS trials, depicted for each of 48 in-network channels across the 3 subjects. **(g)** T-statistics reflecting Post-Pre iTBS change in power for each in-network channel, with values below zero reflecting iTBS-related power decreases. While the numerical mean of these distributions is below zero for both frequency bands, mixed-effects analysis did not reveal a significant change in either. **(h)** Post-iTBS single pulses were binned into sub-blocks of about 12 trials each, similar to Figure 6c. In-network spectral power was measured in the 0.7-0.85 s interval and averaged for each sub-block, to assess change in cortical reactivity over time. Greater suppression of theta and HFA power is observed in earlier time periods closer to the offset of iTBS, though this is not universal across subjects. **(i)** In-network power *t*-statistics were correlated with iTEP amplitude, as done in Figure 3d. An inverse correlation was found between theta *t*-statistics and iTEP amplitude (*p*=0.018 by mixed-effects model), but not HFA (*p*=0.185). Graphic in panel (a) created partly in BioRender.

In an example subject (Figure 8a-d), we observed in-network decreases in both theta and HFA in our pre-specified modulation window, while qualitatively noting more substantial power decreases during the 0-400ms period of elevated broadband spectral power. Across the population of three included participants, average power across channels in the 0.7-0.85 s window numerically decreased from pre to post iTBS in the theta and HFA bands, though we did not detect a subject-level significant effect (theta: *z*=-1.47, *p*=0.142; HFA: *z*=0.518, *p*=0.604 by mixed-effects model). The lack of an HFA effect was driven by one participant, Subject 841, who exhibited a pre-iTBS to post-iTBS increase in power, opposite the other two (Figure 8e-g).

In our TMS experiment, single-pulse blocks began between 3 and 4 minutes following offset of iTBS, raising implications for where in the post-iTBS “recovery” period we might be measuring cortical reactivity. By examining the change in pulse-provoked power over the course of the single-pulse stimulation block (each spanning approximately 125 seconds), we noted increases in theta and HFA power in subject 720, minimal change in subject 789, and increases in HFA in subject 841 with concomitant theta decreases (Figure 8h). Pooling channels across all subjects, we noted that spectral power was maximized at the end of the single-pulse block (approximately 5 minutes after iTBS).

Finally, as noted in our electrical stimulation experiment, we found that iTEP amplitude was inversely correlated with pulse-provoked theta and HFA power, though this relationship was only significant in the theta band (Theta: β=-0.551, *p*=0.018; HFA: β=-0.209, *p*=0.185 by mixed-effects model; Figure 8i).

To address the possibility that TMS-iTBS results in stronger HFA changes in a different modulation window, we expanded our assessment and found significant Post-Pre HFA suppression in the 0.15 to 0.4 s window (Supplemental Figure 8a). In this window, we noted concomitant theta suppression (Supplemental Figure 8b) as well as a significant inverse relationship between iTEP amplitude and Post-Pre change in both HFA and theta power (Theta: β=-0.726, *p*=0.022; HFA: β=-0.653, *p*=0.035; Supplemental Figure 8c). These findings closely mirrored the dynamics observed in our intracranial experiment, but in an earlier window of time that overlaps the evoked potential itself.

## Discussion

Brain stimulation has opened exciting new avenues for basic neuroscience and translational research, but its reach and potential utility has been limited by our poor understanding of how it works. iTBS stands as one of the most widely-used interventions for affecting plasticity, hypothesized to occur by promoting LTP^6,7,9^. However, we do not know whether it truly has this effect in the brain – even foundational studies regarding iTBS to human motor cortex are called into question by more recent and better-powered evidence^13^. To address this gap, we conducted two experiments. First, in 20 neurosurgical patients, we delivered repeated sessions of iTBS intracranially and measured cortical reactivity via single electrical pulses before and after each session. Second, in 3 neurosurgical patients, we delivered a session of iTBS via TMS, measuring cortical reactivity with single pulse TMS. To further generalize our findings, we also examined measures of excitability during resting-state iEEG recordings before and after iTBS.

Across these experiments and analyses, we found a consistent but surprising pattern: within the network of regions closely connected to the stimulation site, the most immediate effect of iTBS was inhibitory. Cortical reactivity – as indexed by spectral power in the HFA and theta bands – was decreased in the immediate aftermath of iTBS. Additionally, stronger effective connectivity to the stimulation site yielded greater iTBS-related suppression. Evidence of this was present in both our intracranial electrical stimulation experiment and our TMS experiment. However, the story grew more complex as we examined the time between sessions of iTBS. As time progressed in the aftermath of iTBS, we noted increased spectral power and cortical reactivity, reflecting the kind of enhanced excitatory activity we might have expected from early studies of iTBS in motor cortex^9^. Though we were not statistically powered to make strong claims about the precise timing of this phenomenon, it was significant about 4-6 minutes after offset of iTBS. We further noted intriguing suggestions of a dose-dependent effect – wherein the strength of suppression and rebound excitation enhances with each iTBS dose, and reaches significance by session 3 – but a linear test for progression across blocks was subthreshold for significance.

This suppression-rebound dynamic was not observed in brain regions with weak connections to the stimulation site (out-of-network), reinforcing that this is not a generalized, brain-wide reaction to stimulation, but rather a network-specific phenomenon. While sham periods served as a useful control and demonstrated no statistically significant effect on average (Figure 2c), we note that our experimental design – which alternated between active and sham sessions – effectively positioned “sham” periods during the iTBS recovery. As such, we noted the appearance of a sham-related upregulation in excitability that we attributed to an ongoing post-iTBS recovery. This raises an important experimental design challenge for future work, especially in time-constrained settings wherein active and sham interventions are spaced close together.

Our results suggest a dynamic neural response to iTBS. There may be an initial “refractory” period – in our experiment around 1 minute – during which time cortical reactivity is relatively suppressed. However, the stimulated network rebounds to elevated excitatory drive as time passes, becoming the predominating effect. Critically, this was observable owing to our experimental design, wherein CCEPs begin immediately upon offset of iTBS. The motif of delayed stimulation-induced excitation has been noted across prior *in vitro* and *in vivo* studies^44–47^, though the observed timescale ranges from minutes to hours. A minutes-long progressive increase in excitability, as we observed here in human neocortex, might reflect activity-dependent functional synaptic changes leading to enhanced recruitment of neurons downstream of the stimulation site^48^. This timescale is too short to classify as LTP itself and likely reflects a more acute form of plasticity, potentially correlated with longer-term changes that will be explored in future experimentation.

Our TMS-iTBS experiment also showed evidence of rising cortical reactivity over time, noting that the subject with the greatest cortical reactivity (841) began spTMS the latest after iTBS (3.7 minutes), while the subject with the most suppression (720) began the earliest (3.3 minutes). As we utilized the same modulation window (0.7-0.85 s) to measure TMS-related effects, we consider these results a conservative test of generalization; the underlying physics of TMS and intracranial electrical stimulation differ greatly^38,40,49^ and may not be expected to alter plasticity in the same way. As shown in supplementary analyses, significant population-level HFA modulation was observed in our TMS experiment at earlier timepoints (0.15-0.4s) than electrical stimulation, suggesting either (1) the directionality of effect is consistent across TMS and direct electrical stimulation, even as the precise timing of effects differs, or (2) a larger TMS sample would reveal a closer match in the timing properties of iTBS-related changes in cortical reactivity.

Evidence of a suppression-rebound following iTBS is further supported by our resting-state analysis, which concerns non-stimulated periods of rest that occur several minutes following iTBS offset. In these analyses, we found that iTBS increased a spectral measure of excitation^35,50,51^, even in the absence of pulse-provoked activity. Our finding of immediate pulse-provoked inhibition following iTBS, but resting-state excitation, may at first seem contradictory. However, our measure of resting-state excitability took place after the full block of post-iTBS single pulses (including an un-analyzed block of low-amplitude pulses, see *Methods*), lasting a total of 3 minutes. As such, we anticipated these measures would reflect the post-iTBS rebound excitation phase, and our observation of flatter aperiodic slopes in the post-iTBS period was consistent with this expectation. We also noted that, at other times, in-network regions tended to exhibit higher exponents and were therefore in a more inhibitory state than out-of-network regions; this could reflect a general property of regions that tend to have high effective connectivity with the DLPFC (which dominated stimulation sites in that subset of subjects), or potentially an effect of CCEPs acting as a form of low-frequency repetitive stimulation.

Our findings could help explain some of the variability present in the literature on iTBS-related effects in human cortex^22,52^. Experimental designs vary when they probe for cortical reactivity following iTBS, and this timing may matter. Moreover, some protocols involve extended periods of single-pulse probes, ranging to 20 minutes or beyond – in these cases, it would be important to not analytically treat these probes as “one block,” but rather examine subdivisions that could stratify effects over time. Some prior studies and meta analyses show evidence for time-related effects in measures of motor-evoked potentials^11,13^, potentially reflecting underlying iTBS-rebound phenomena that were not fully accounted for in the original experimental design. Finally, we showed evidence that iTBS effects may accumulate over repeated sessions, suggesting single-dose experiments may not find evidence of excitation at all. Robust effects – particularly when measured with non-invasive tools like scalp EEG – may require many iTBS sessions to reliably manifest^53^, paralleling symptomatic change in multi-day clinical iTBS protocols.

We avoided detailed analysis of region-specific effects of iTBS, for several reasons. First, considering all in-network channels regardless of stimulation site (or recording site) allows us to make a more general claim about the effect of iTBS on cortical function, extending even beyond the DLPFC. However, even if we adopted a more liberal threshold for in-network inclusion, we may be underpowered to make strong claims about subregional effects without collecting more data. Future work should examine whether iTBS effects differ depending on the cortical site selected, as well as other anatomical factors (e.g. proximity to white matter, gray matter, or the gray/white boundary).

To constrain our scope and to address a focused hypothesis regarding the aftereffects of iTBS, we did not analyze neural activity that occurs during iTBS itself. However, the dynamics that unfold during stimulation may provide further clues to the mechanism underlying plasticity-modulating interventions in the brain, and should be the focus of future work. In particular, there may be spectral signatures that occur during iTBS or the 8-second inter-train interval which correlate with subsequent post-iTBS effects such as those analyzed here, providing a more unified account of how iTBS exerts its neuroplastic effects.

This study is a meaningful advance in our understanding of patterned brain stimulation protocols, demonstrating a dynamic process of iTBS-related suppression of reactivity with rebound excitation in human neocortex. While the story is far from complete, these data can guide future investigations into plasticity protocols by (1) emphasizing the need for standardized experimental procedures, down to minute-level timing, and (2) highlighting how effects of iTBS can vary over time. This work lays the foundation for studies that will examine how neural plasticity is influenced by stimulation parameters, offering insight into the fundamental mechanisms of LTP and excitatory/inhibitory balance in the human brain.

## Methods

### Human subjects: Electrical stimulation/iEEG

From 2023-2026, 20 neurosurgical patients with medically intractable epilepsy underwent a surgical procedure at Stanford University Hospital to implant intracranial recording contacts within brain parenchyma (stereo-EEG). Contacts were placed in accordance with clinical need to localize epileptic regions. All patients gave their written informed consent for the surgical procedure and to participate in brain stimulation research during their hospital stay. Stimulation experiments were conducted shortly before electrode explantation, after patients had been re-loaded on antiepileptic medications. Research was approved by the Stanford University IRB.

### Human subjects: TMS/iEEG

Three neurosurgical patients, also with medically intractable epilepsy, underwent a surgical procedure at the University of Iowa Hospital and Clinics to implant intracranial recording contacts within brain parenchyma. Contacts were placed in accordance with clinical need to localize epileptic regions. TMS experiments were conducted after the final surgical treatment plan was agreed upon between the clinical team and the patient, typically 1-2 days before the planned electrode explantation operation and 24 hours after the patient had been re-loaded on antiepileptic medications. Research was approved by the University of Iowa IRB and written informed consent was obtained from all participants.

### Imaging protocol and intracranial electrode localization

For each patient, post-implant CT images were coregistered with presurgical T1-weighted MRI using FLIRT from the FMRIB Software Library (FSL)^54–56^. Surface reconstruction was generated from the T1 scan using the recon-all command of Freesurfer v7.3.2^57^. Each electrode on the T1-registered CT was identified using BioImageSuite^58^. Electrode coordinates in the FreeSurfer surface space, voxel space, and MNI305 space were automatically extracted by the iElVis toolbox^59^, which were then labeled according to their location within the Desikan-Killiany-Tourville (DKT) anatomical atlas.

### Stimulation protocol: Electrical stimulation

All single-pulse stimulation (spES) was delivered through bipolar channel pairs (0.2 ms pulse duration) as 7mA biphasic square waves, using a Nihon Koden electrical stimulator. For most single-pulse stimulation blocks, we delivered 27 pulses at 0.3 Hz (lasting 90 seconds; see Figure 1a-b). Single-pulse blocks also included another set of 27 pulses at 0.3 Hz delivered at 4mA, but these were not analyzed in the present manuscript. iTBS was delivered at 4mA, comprising a session of 20 trains of 50 Hz triplets spaced 200 ms apart, delivered for 2 seconds on and 8 seconds off (total duration of 600 pulses delivered over 3 minutes). Fourteen subjects underwent an experiment wherein single pulse blocks were delivered before and after 3 iTBS sessions, preceding each iTBS block by a 1.5 minute delay and starting immediately after offset of iTBS. Three sham blocks, in which no stimulation was given for 3 minutes, were interleaved with iTBS blocks as depicted in Figure 1b. Six subjects underwent an experiment wherein we delivered one session of iTBS, one session of Sham, and one session of continuous TBS (cTBS) that was not analyzed in this manuscript, and as such these subjects only contribute to a measurement of one block of iTBS (“iTBS1”). In this 1-session iTBS study, single pulses were delivered at 0.4 Hz for a total of 48 pulses per block. Note that, due to clinical needs and other constraints of conducting experiments in the hospital setting, modest deviations from protocol timing were tolerated, and subjects were allowed to discontinue testing prior to completion of our full protocol should clinical needs arise; 8 subjects completed the entire 3-session protocol with no major interruption or deviation, while 6 experienced some interruption or abridgment of at least one single pulse or iTBS block.

For the 14 participants who were assigned to the 3 session iTBS study, only one stimulation site was selected; the 6 who completed the 1 session iTBS study could receive stimulation at up to 3 unique sites. Stimulation targets bilaterally spanned several cortical and subcortical sites, including dorsolateral prefrontal cortex, posterior parietal cortex, cingulate gyrus, and amygdala (see Supplemental Table 1). Seizure onset zones or regions exhibiting frequent inter-ictal activity were strictly avoided as stimulation sites.

### Stimulation protocol: Transcranial magnetic stimulation (TMS)

Stimulation was delivered using a MagVenture MagVita X100 230 V system with a figure-of-eight liquid-cooled Cool-B65 A/P coil (MagVenture, Alpharetta, GA, USA), automatically controlled via the BEST toolbox^60^. Stimulation pulses were biphasic sinusoidals with a pulse width of 290 us, with stimulator output set at a percentage of each subject’s motor threshold (see below). All TMS experiments were conducted after re-starting antiepileptic medications. Neuronavigation using frameless stereotaxy was guided with Brainsight software supplied with the pre-implantation T1/MPRAGE scan. Stimulation parameters were recorded in Brainsight during all experimental trials. Motor thresholds were determined starting with the hand knob of the motor cortex as a target, beginning at 50% machine output and adjusted in 2-5% increments until movements of the intrinsic hand muscles were observed in 50% of trials.

Single pulses and iTBS were directed at DLPFC targets at or above the motor threshold (100% was used if 120% was not tolerated due to pain). Targets were defined by the Beam F3 region^61^, identified by transforming published coordinates (MNI 1mm: -41.5, 41.1, 33.4)^62^ into each subject’s native T1 and displaying it in Brainsight. Single pulse TMS was delivered with jittered 2.5 s inter-stimulus intervals. Blocks of 50 single pulses (spTMS) were delivered before and after sessions of iTBS. Pre-iTBS single pulses occurred between 3-5 minutes prior to iTBS, while post-iTBS single pulses occurred between 3-4 minutes after iTBS offset (see Figure 8h). In two subjects (789 and 841) spTMS and iTBS were delivered at 120% of motor threshold; subject 720 underwent 102% for spTMS and 82% for iTBS (see *Suppression-rebound following direct electrical iTBS generalizes to TMS-iTBS*).

### iEEG recording and preprocessing

AdTech Medical (Racine, WI, USA) electrodes were used for experiments at both Stanford University Hospital (electrical stimulation) and the University of Iowa Hospital and Clinics (TMS-iEEG). At Stanford, data were recorded on a Nihon Koden system at 1000 Hz, while at University of Iowa a NeuraLynx (Bozeman, MT, USA) ATLAS Neurophysiology system was used. In all subjects, contacts were excluded from analysis if they were determined to be involved in the generation or early propagation of seizures, or if electrodes were contaminated by nonneural noise indicative of poor connection or placement outside the brain. All recording contacts on the same lead (i.e. stereo electrode shaft) as the stimulated electrode were excluded from all analyses.

Data preprocessing and analysis was principally carried out using the MNE Python toolbox^63^. To account for large-scale noise or contamination of the reference electrodes, signals were re-referenced using a bipolar montage. We avoided running inferential statistics on time periods contaminated with artifact from stimulation pulses, but it was necessary to spectrally decompose these periods for the purpose of generating time-frequency representations and visualizations (e.g. Figure 1f, Figure 4). As such, we adopted a procedure to scrub stimulation artifact from all signals and replace it with synthesized stationary iEEG that reflects a similar spectral profile as the background^43,64^. Specifically, iEEG signal was clipped from 15 ms prior to 15 ms following each stimulation pulse and replaced with a weighted average of the 30 ms immediately following and prior to stimulation. Pre- and post-stimulation clips were first reversed, then tapered linearly to zero along the length of the signal, and then finally summed together to replace the artifact period. Finally, signals were notch filtered at 60 Hz and harmonics to remove line noise, using an F-test to find and remove sinusoidal components^65^. Signals were then downsampled to 500 Hz for further analysis.

To remove iEEG signal that may have been contaminated by epileptiform spikes or other artifactual transients, signals were *z*-scored across all trials (see *Stimulation protocol: Electrical stimulation*) and timepoints on a channel-level basis, and any trial in which one or more samples exceeded an absolute *z-*score threshold of 5 were discarded from further analysis. Any channels for which more than 7 trials were discarded (leaving at a minimum 20 trials left for analysis) were themselves marked as “bad” and removed from further analysis.

### CCEP/iTEP analysis

To generate the cortico-cortical evoked potentials (CCEP) or intracranial TMS-evoked potential (iTEP), we adopted a standard approach wherein the voltage trace for each trial was first baseline corrected (*z-*scored) to the -200 ms to 0 ms window, then averaged across all trials which occurred before the first iTBS session (Figure 1c-d). The peak-to-peak (PTP) amplitude of the CCEP/iTEP was used as an index of effective connectivity with the stimulation site (prior to any neuroplastic intervention), calculated by taking the maximum-to-minimum value within the 50 ms to 300 ms interval (reflecting the N2 component of a canonical CCEP). This PTP value was then used as the basis for thresholding in-network vs out-of-network channels; most of this manuscript presents data for a threshold of 10, though robustness of observed effects at lower thresholds is also demonstrated (see Figure 3a-b, and Supplemental Figure 1, Supplemental Figure 2, Supplemental Figure 4).

As a control, we also examined iTBS-related effects in a group of “out-of-network” channels that demonstrated poor effective connectivity with the stimulation site (Figure 2a). To identify this subset, all channels in each subject were ranked according to their CCEP PTP amplitude, as outlined above. For each subject, if *n* channels were identified as in-network (i.e. under the >10 threshold for analyses presented in the main text), a corresponding group of *n* lowest ranked channels were selected as out-of-network; this procedure matches the number of channels in overall number while selecting for those that demonstrated poor responses to single-pulse stimulation.

### Spectral analysis

To compare pulse-provoked spectral activity between pre- and post-iTBS blocks of single pulse events (“trial”), iEEG signals from each channel (reflecting a bipolar pair of recording contacts) were epoched into 2.5 s intervals, spanning 500 ms prior to a pulse until 2 s after. To decompose the signal into component frequencies, we used the multitaper method (time-bandwidth product of 4, excluding tapers with <90% spectral concentration). For our initial HFA analyses (Figure 2), we used sliding windows of 6 cycles in length at 6 log-spaced frequencies from 75 Hz to 180 Hz, averaged the result across frequencies, and then subtracted the pre-pulse baseline power averaged in the - 600 ms to -100 ms window. Finally, the resulting baseline-corrected timeseries was temporally averaged into 200 ms windows (spaced 50 ms apart) to generate our final HFA timeseries (see Figure 1f, Figure 2b). Theta-specific analyses were conducted similarly, but instead using windows of 2 cycles in length for 4 log-spaced frequencies from 4 Hz to 8 Hz and baseline corrected to the -750 ms to -250 ms window. For the full-spectrum analysis presented in Figures 4, 6, and 8 we used 35 log-spaced frequencies from 4 Hz to 180 Hz, dynamically scaling the number of cycles per frequency from 2 to 6.

### Statistical approach: Cortical reactivity

As our central hypothesis concerns the iTBS-related change in spES-provoked spectral power (termed “cortical reactivity”) from pre- to post-iTBS, we used a 2-sample *t-*test to compute the post-minus-pre change in spectral power for each channel at each 200 ms time bin, comparing the “post” distribution to the “pre” distribution of power values (see *Spectral analysis* and Figure 1f-g). Trials are not independent measurements, and therefore the *p*-values from this test would be inflated and were not considered, nor was any statistical inference made from this test directly. A *t*-statistic was generated for each channel and each time bin, reflecting whether power in a given band increased or decreased from pre- to post-iTBS (negative values indicate iTBS-related decreases in spectral power, see Figure 1g and Figure 3a-b). These *t*-statistics were used as the basis of subsequent statistical tests as described below. The number of subjects included and mixed effects model specifications for all key tests are summarized in Supplemental Table 2.

#### Population-level HFA effect (Figure 2b-d)

As placement of electrodes in neurosurgical patients is dictated by clinical need, subjects had widely varying numbers of recording contacts that were ultimately labeled as in-network (see Supplemental Table 1). Additionally, due to constraints of working in the hospital environment, subjects completed variable quantities of the experiment (see *Stimulation protocol: Electrical stimulation*). To account for this variability, the presence of missing data, and the hierarchical nature of the data (wherein effects at individual recording channels are observed within subjects), we adopted a linear mixed-effects model approach for major statistical analyses in this manuscript^66^. Specifically, using the Python statsmodels package^67^, we first used an intercept-only model to test for a significant population-level effect of iTBS across all subjects, channels, and experimental sessions in the HFA band. For each 200 ms bin, the channel-level *t*-statistic was modeled as a function of iTBS block (fixed effect) and subject (random effect). The Wald test was used to ask whether the intercept significantly deviated from zero, providing a *p-*value and Wald *z*-statistic for each 200 ms bin reflecting whether there is a population-level effect of iTBS (Figure 2b). *P*-values were FDR corrected for multiple comparisons across time bins, and bins with a corrected value of *p*<0.05 were taken as a focus window for other analyses in the manuscript. (Of note, models did not converge for time bins between 1.0 and 1.25 s, so effects were approximated with 1-sample *t-*tests across subjects after averaging effects across channels; none of these time bins were significant.) The same approach was taken to ask whether sham intervals yielded a significant change in HFA at any time bin (Figure 2c), or whether changes were observed in the out-of-network group of channels (Figure 2d). After the significant window of HFA modulation was identified, a combined model was used to test for significant differences between effects in active vs. sham and in-network vs. out-of-network (block, network category, and active vs. sham as fixed effects, subject as random effect).

#### HFA vs. CCEP correlation (Figure 3c-d)

To ask whether there was a correlation between CCEP amplitude (see *CCEP/iTEP analysis*) and channel-level HFA t-statistics (Figure 3a-b), we specified a model wherein channel *t*-stats were a function of PTP amplitude (fixed effect), Euclidean distance (fixed effect), and subject (random effect). *T*-stats were first averaged across all iTBS blocks per channel, so block was not included as a factor in this model. Random slopes with respect to PTP and distance were fit per-subject. Prior to fitting the model, PTP and HFA *t*-stats were re-scaled by first centering the values within subjects, and then dividing by the pooled standard deviation across all channels/subjects (after centering). Pearson coefficients reflecting within-subject correlations are shown for two example subjects in Figure 3c and Supplemental Figure 3, but these are solely descriptive and not used to make population-level inferences.

#### Frequency band analysis (Figure 4)

Tests for significant iTBS-related modulation of spectral power in several frequency bands were conducted similarly to the HFA tests described above. Specifically, channel-level *t*-statistics were computed for each time bin and each frequency band of interest, spanning theta (4-8 Hz), alpha (9-13 Hz), beta (15-25 Hz), gamma (30-55 Hz), as well as HFA as previously computed. As with HFA, intercept-only models were used to model channel-level *t*-statistics as a function of iTBS block (fixed effect) and subject (random effect), and the resulting *p*-values were then corrected for multiple comparisons (Bonferroni) across the 5 tested frequency bands, in the previously-established window of interest (0.7-0.85 s).

To assess for a linear progression of iTBS effect across blocks (Figure 4c), for each trial spectral power was averaged in the window of interest established earlier (0.7-0.85 s), and the average power of all “pre” blocks was subtracted from the average power of all “post” blocks. This procedure was repeated for each iTBS session individually, generating a measure of Post-Pre change in power for each session and each channel. A mixed effects model was constructed to predict this power difference as a function of ordered iTBS block number (fixed effect) and subject (random effect).

#### Active vs. Sham and Out-of-Network analyses (Figure 5)

Intercept-only mixed effects models were used to assess for significant power modulation at each individual iTBS block. Channel-level *t*-statistics were modeled with subject as a random effect for each sham session and active iTBS session individually. This procedure was repeated for the group of out-of-network channels. Across the three iTBS blocks, *p*-values were Bonferroni corrected and considered significant at the level of *p*<0.01.

#### Post-Active vs. Post-Sham change in spectral power (Figure 6)

To ask whether spectral power changed in the time periods between iTBS sessions, suggesting a possible rebound effect, we examined cortical reactivity changes between the Post-Active block (immediately following an iTBS session) and the Post-Sham block (immediately following the next 3-minute Sham session). This provides an approximately 15-minute window of time over which subjects undergo spES and rest periods, but no iTBS. Our statistical approach was similar to our evaluation of pre- vs post-iTBS power changes. Specifically, for each in-network channel, spectral power was averaged across all Post-Active trials and Post-Sham trials, and then subtracted from one another (Post-Sham minus Post-Active) to generate a measure of power difference over this time period. These difference values were modeled as a function of block (fixed effect) and subject (random effect), to estimate the population-level effect across all blocks. Each block was also tested individually as an intercept-only model with subject as random effect; the resulting *p*-values were Bonferroni corrected and considered significant at the *p*<0.01 level (Figure 6b).

In a subset of 8 subjects who completed all three iTBS blocks with limited deviation from our canonical protocol timing, we further subdivided each block of spES into three sub-blocks of 9 trials each (Figure 6c), to get a more precise measurement of cortical reactivity rebound effects. Within the previously-identified 0.7-0.85s window, HFA power (corrected to pre-pulse baseline) was averaged in these 9-trial bins and plotted with respect to elapsed time in the experiment; Figure 6c shows average elapsed time across the 8 subjects included in this analysis. Intercept-only models (modeling averaged HFA power as a function of subject-level random effect) were used to estimate whether power was significantly above or below baseline for each spES block.

#### TMS-iTBS experiment (Figure 8)

Our statistical approach to evaluating for Post-Pre TMS-iTBS change in cortical reactivity mirrored our procedures outlined above, with the notable modification that each subject only underwent one iTBS session. Intercept-only mixed effects models were used to test for a population-level effect in each frequency band, with subject as a random effect (Figure 8d-e). Correlations between power *t-*statistics were assessed via a mixed model specifying subject (random effect) and iTEP amplitude (fixed effect), with random slopes fit per-subject. Prior to fitting the model, values are re-scaled by first centering the values within subjects, and then dividing by the pooled standard deviation across all channels/subjects (after centering).

### Statistical approach: Resting state excitability

To ask whether measures of cortical excitability during resting state were changed by iTBS, we examined the exponent of the power spectrum, which is believed to reflect greater inhibition at steeper slopes and greater excitation at shallower slopes^35,36^. To measure the spectrum exponent, each resting state period (comprising non-stimulated periods of time between single pulse blocks and iTBS blocks as well as Sham periods, see Figure 1b) was divided into consecutive 3-second epochs of time. Any block with less than 30 seconds of total resting time was not included in the analysis. For each 3-second period we used the FOOOF toolbox in Python^36^ to extract an estimate of the oscillatory and aperiodic components of the power spectrum (peak width limits = [0.5, 12.0], maximum peaks = 4, minimum peak height = 0.1, peak threshold = 3.0, fixed aperiodic mode). Spectra were measured using the multitaper method as described previously, from 3-180 Hz. This procedure provided an estimate of the spectrum exponent for each 3-second epoch spanning all resting state periods of the experiment. Of note, we performed this analysis in N=9 subjects who not only met channel inclusion threshold but also included sufficient resting state time and who completed the version of the experiment with more than 1 iTBS session (see *Stimulation protocol: Electrical stimulation*).

Next, exponents were *z*-scored to the mean and standard deviation of each subject’s first resting state block of the experiment. For each channel, *z*-scored exponents were averaged across all epochs within each resting-state block. The difference in averaged exponent was taken between the resting state block most closely preceding and following each iTBS session (Figure 7a); of note, a 1.5 minute period of rest immediately precedes each session, but each session is immediately followed by a block of CCEPs prior to the post-iTBS resting period. As such, exponents estimated post-iTBS are measured after a 3-minute delay from iTBS offset. Channel-wise difference scores were modeled using a similar mixed-effects approach as outlined previously, specifying subjects (random effect), iTBS block (fixed effect), and in-network versus out-of-network groups (fixed effect). Tests for effects at the level of each individual iTBS session were performed using paired *t*-tests, as small subject Ns caused convergence errors in the mixed-effects approach. 6 subjects were included for iTBS1, 6 for iTBS2, and 7 for iTBS3; variability was driven by resting state periods that were shortened or interrupted, precluding measurement of a full 30 seconds or greater. To generate the exponent timeseries as depicted in Figure 7c, the average *z*-scored exponent was plotted for every resting state block in the experiment.

Correlations between HFA change and exponent change (Figure 7d) or CCEP amplitude and exponent change (Supplemental Figure 6) were computed as follows. For the iTBS3 block only – where Post-Pre exponent change was determined to be significant – all channels with CCEP amplitude >3 were taken (as done in Figure 2d and further evaluated in Supplemental Figure 4). A mixed model was fit to evaluate the relationship between HFA change (average *t*-stat in the 0.7-0.85 s window) and Post-Pre exponent change, specifying subject as a random effect and fitting per-subject slopes with respect to HFA change. The same statistical approach was taken to model the relationship between CCEP amplitude and Post-Pre exponent change.

## Funding

NIMH R01MH132074 and NIMH 5R01MH139650 (CJK, NTT, and ADB); R21MH120441, R01NS114405 and Roy J. Carver Trust (ADB); R01MH126639, R01MH129018, and Burroughs Welcome Fund Career Award for Medical Scientists (CJK); 5T32-MH019113, NIMH 1K23MH125145, NIMH 5R01MH136197, BBRF Young Investigator Grant 31275, and the Penningroth Fellowship in Interventional Psychiatry (NTT); T32MH019938 (CWD and EAS); K99MH141192 and the Sleep Research Society Foundation CDA 049-JP-26 (UH); and 5K99MH139907 (EAS). This work was conducted, in part, on an MRI instrument funded by 1S10OD025025-01.

### Acknowledgements

We thank Josef Parvizi his contribution to patient recruitment, imaging analysis, and facilitation of time with the neurosurgical patients at Stanford University. We also thank Joel Berger, Chris Garcia, Ariane Rhone, Kirill Nourski, Joel Bruss, Benjamin Pace, Haiming Chen, Hiroto Kawasaki for their contributions to imaging acquisition and data collection efforts at the University of Iowa. We also thank the neurosurgical patients who selflessly participated in this research. Graphics in panel 1a and 8a created in part in BioRender (Solomon, E. 2026 https://BioRender.com/hxp1e13).

## Data Availability

Upon publication, raw deidentified data will be made available in the Stanford Digital Repository (https://sdr.stanford.edu/). Analysis code is available on a GitHub repository (https://github.com/PrecisionNeuroLab/Analysis-SolomonEtAl2026/).

