## Supplemental Figures for "iTBS induces a suppression-rebound pattern in human cortical excitability"

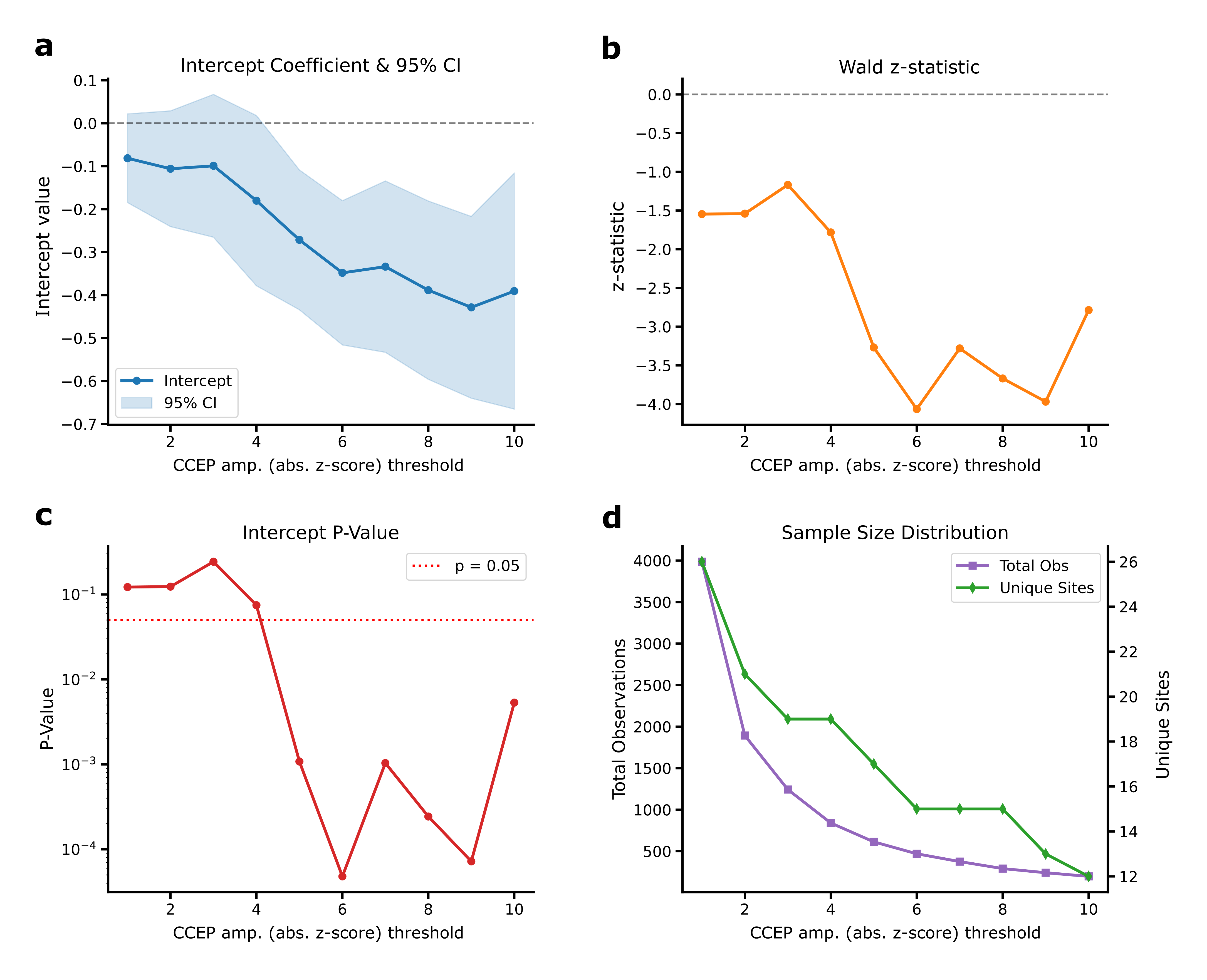

**Supplemental Figure 1. Post-iTBS decrease in cortical reactivity is preserved across CCEP amplitude inclusion thresholds. (A)** Intercept value in an intercept-only linear mixed effects model (see *Methods*) that reflects the average Post-Pre iTBS *t*-statistic in the population, as a function of channel inclusion threshold from 1-10. Negative values reflect HFA decreases from pre to post-iTBS, in the 0.7-0.85 s window identified previously (see Figure 2). Error bars show 95% CI. **(B)** Wald *z-*statistic as a function of channel inclusion threshold. **(C)** *P*-value, reflecting whether the average intercept significantly deviates from zero, as a function of inclusion threshold. Significance (*p*<0.05) is achieved above a CCEP amplitude cutoff of 5 (absolute z-score units). **(D)** Total observations (channel count; left axis) and unique stimulation sites (right axis) as a function of inclusion threshold. Note that in the population of N=20 subjects, several have more than one stimulation site (multiple stimulation sites within a single subject were counted as a virtual “unique” subject in this figure), leading to more observations than the total number of physical channels.

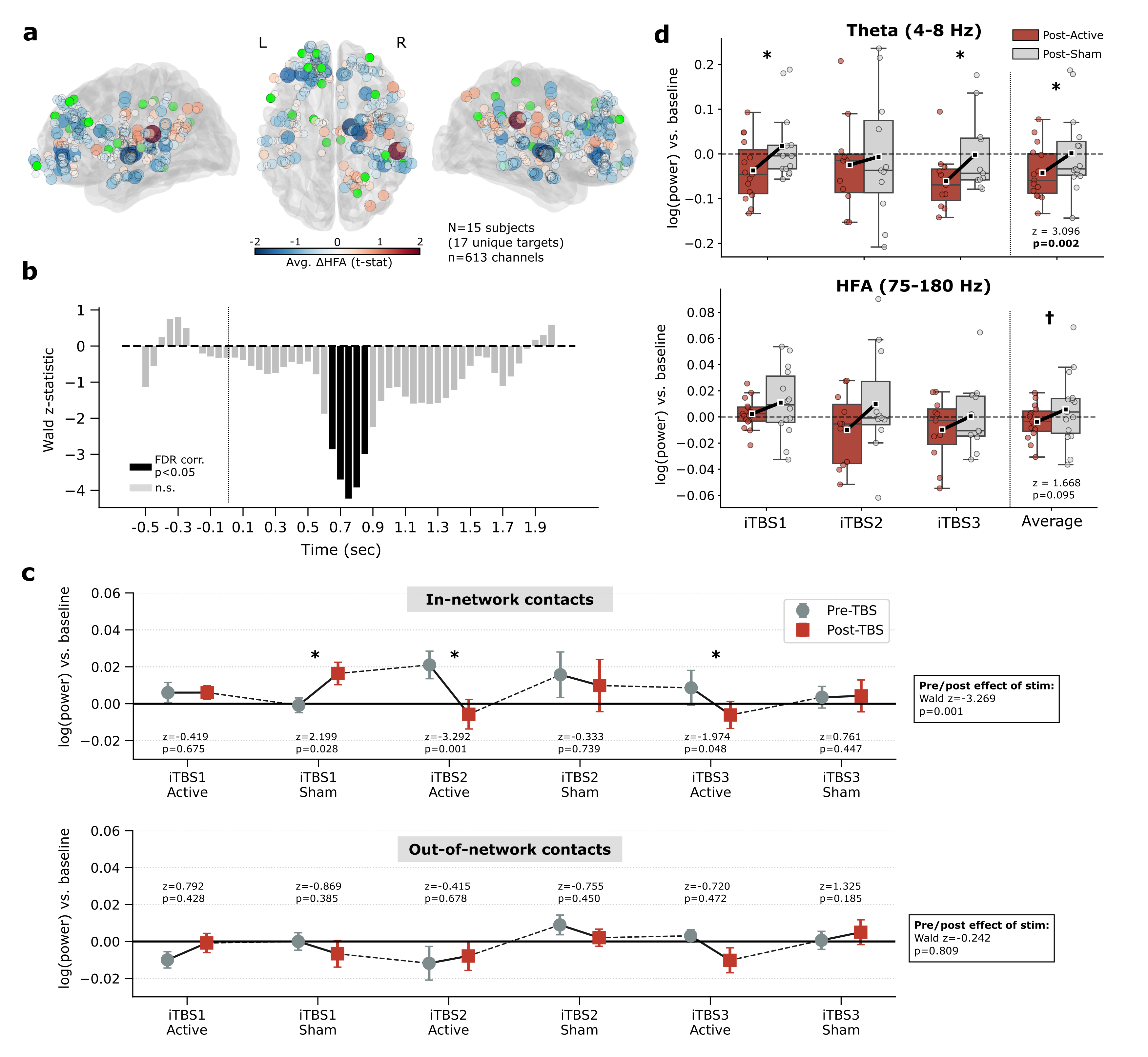

**Supplemental Figure 2. Effects of iTBS are consistent under more inclusive in-network CCEP threshold of >5. (A)** The set of 613 channels across 15 subjects included with a CCEP amplitude threshold of 5, as shown also in Figure 3a. Colors reflect HFA change from pre- to post-iTBS in the 0.7-0.85 s window. **(B)** Population-level ΔHFA effect (Post-Pre) as reflected by the Wald *z*-statistic derived from a mixed effects model, in the 613-channel group. Effects are similar as in the more restrictive 71 channel group (see Figure 2), with significant HFA decreases observed from 0.65-0.85 s. **(C)** *Top:* In the 613-channel group, significant Post-Pre HFA decreases are observed at iTBS2 and iTBS3, with an average effect across all blocks of *t*=-3.269, *p*=0.001. A significant HFA increase was observed for the first sham block (“iTBS1 Sham”). *Bottom:* No effect is observed at any active iTBS or sham block in a sample of 613 out-of-network channels with low CCEP amplitudes. 15 subjects contributed data to iTBS1, 11 subjects contributed to iTBS2-3. **(D)** Replicating the effect shown in main text Figure 6, the 613-channel group demonstrates enhancement of theta and HFA power from the Post-Active to the Post-Sham blocks, though this is a trend-level effect as averaged across all blocks in the HFA band (†*p*<0.1).

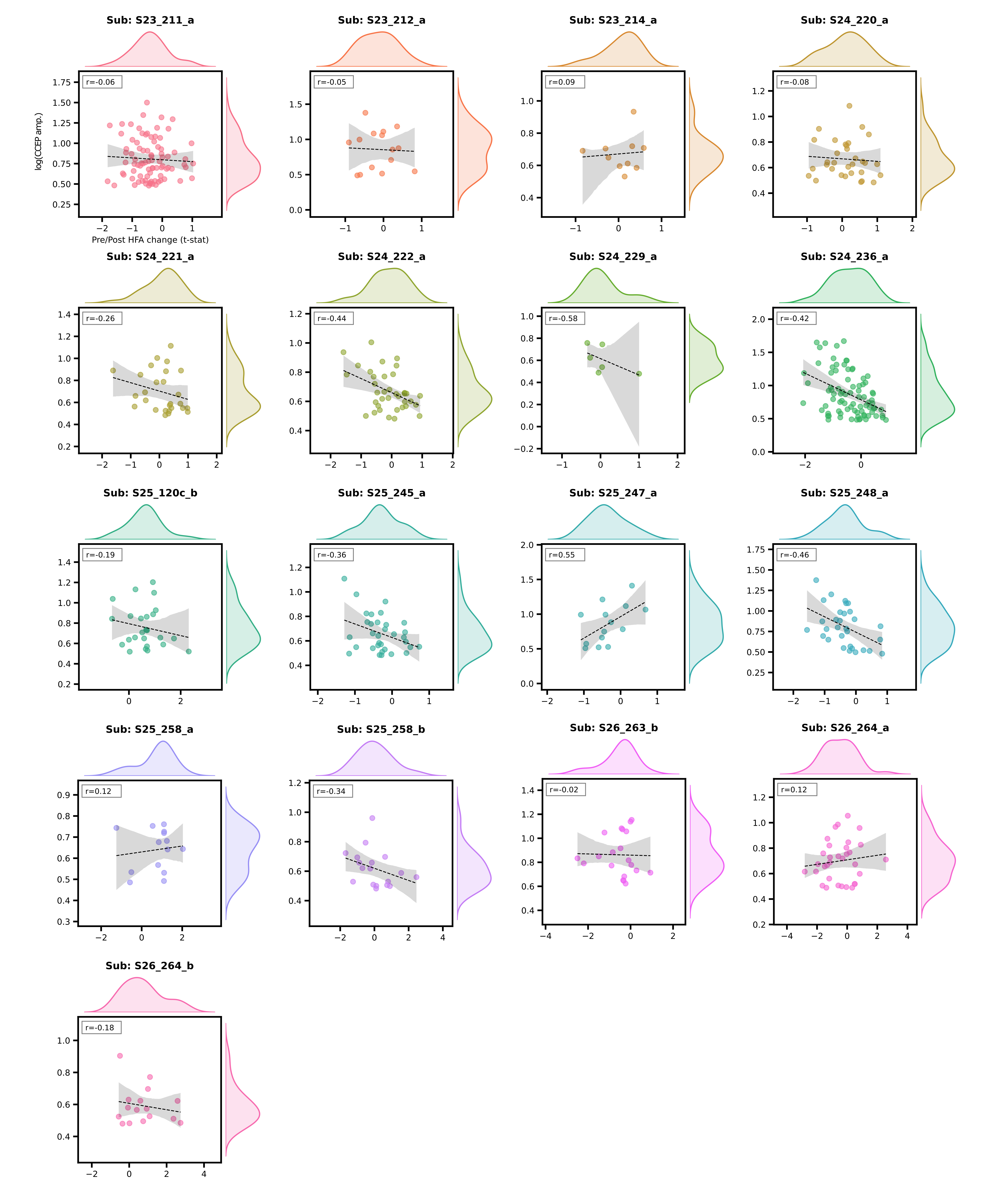

**Supplemental Figure 3. iTBS-related change in cortical reactivity is inversely correlated with CCEP amplitude.** Structured as in Figure 3c, depicted for every subject in the dataset with sufficient data at the CCEP >3 threshold to estimate a correlation (at least 5 channels). Note that subject S26_264 is included under two different stimulation sites. Data reflect Post-Pre HFA power change in the 0.7-0.85 s interval for each channel, versus CCEP amplitude (*z* units).

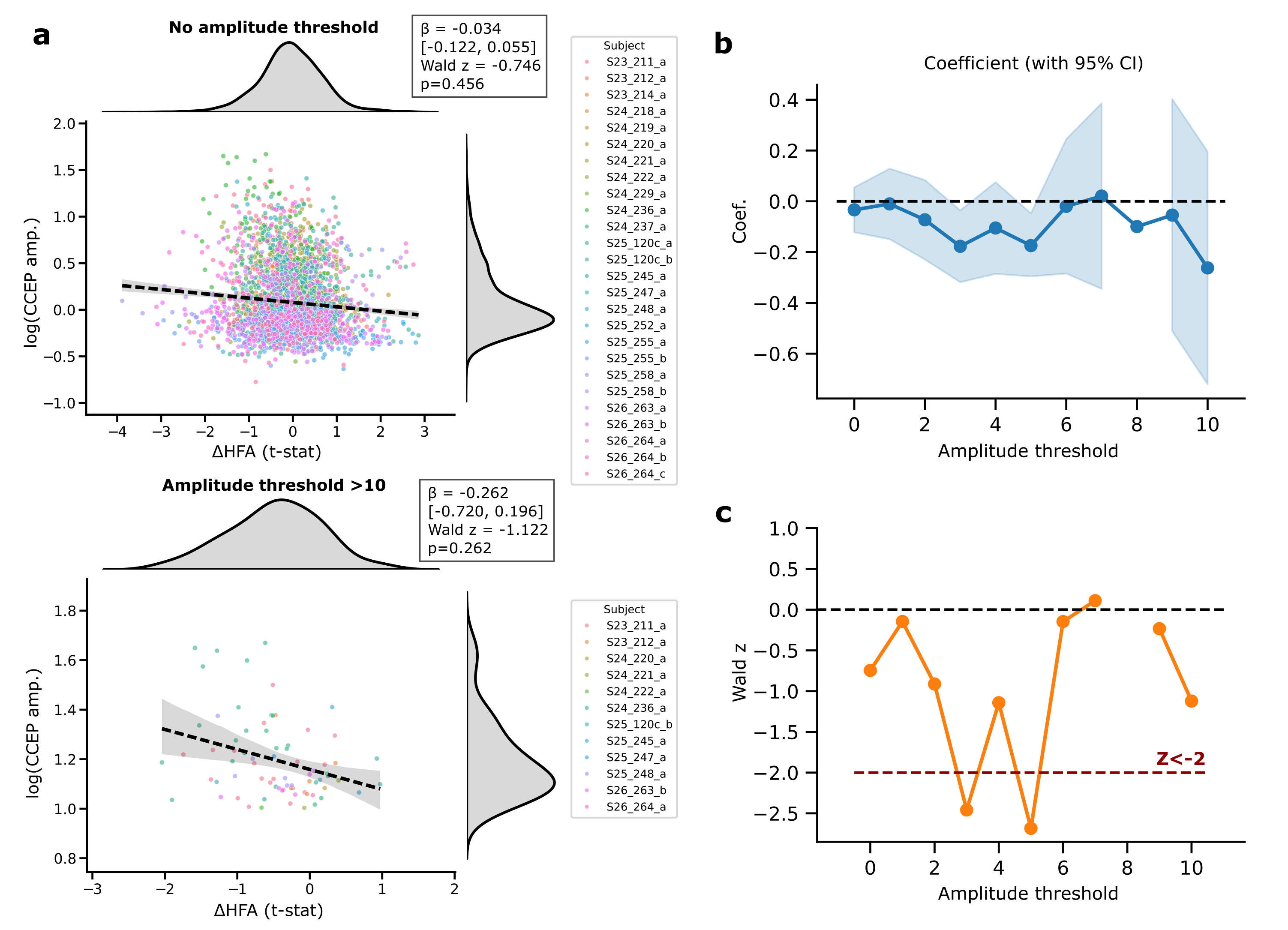

**Supplemental Figure 4. Inverse relationship between change in cortical reactivity and CCEP amplitude is preserved across channel inclusion thresholds. (A)** *Top:* Structured identically to Figure 3d, but with no channel inclusion threshold applied. Across all channels in the entire dataset (n=2772), there is a negative correlation between HFA change in the 0.7-0.85 s interval and CCEP amplitude, but this does not reach statistical significance (see box for mixed effects model output). *Bottom*: An inverse relationship was also preserved using the most stringent CCEP amplitude threshold of >10, though also not meeting significance threshold. **(B)** Coefficients derived from the analysis in (A), depicted for every channel inclusion threshold from 0 to 10. Shaded region depicts 95% CI. Model convergence failure occurred at threshold 8, where the plotted coefficient was instead estimated via Pearson correlation. **(C)** Corresponding Wald *z* values for coefficients estimated at each channel inclusion threshold. Significant (*z*<-2) inverse correlations are observed at threshold of 3 and 5.

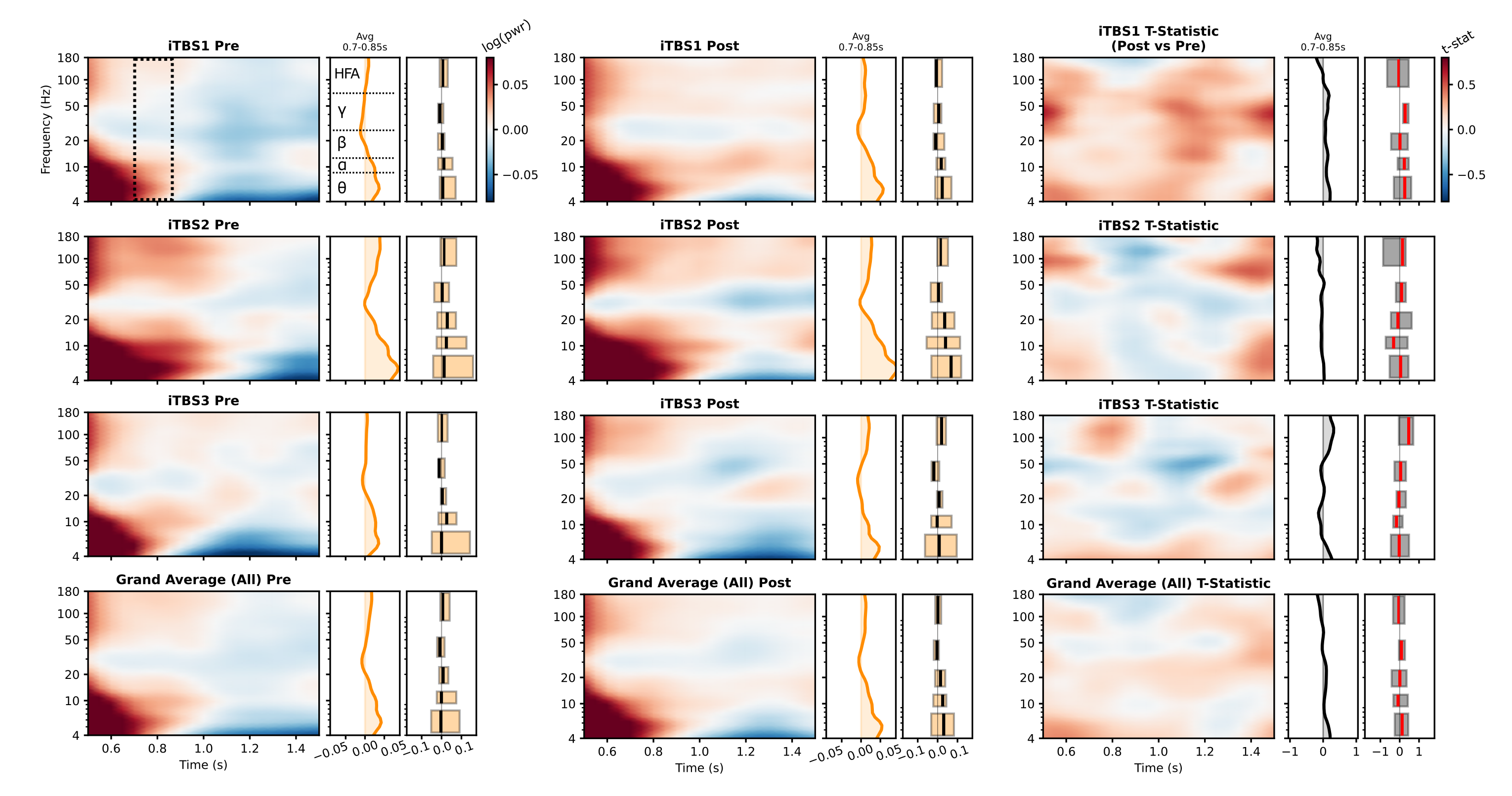

**Supplemental Figure 5. Time-frequency representations for sham-related iTBS effects on cortical reactivity.** Structured identically to main text Figure 4, but for pre-sham and post-sham spES trials. Sidecar plots show the average spectral power within the 0.7-0.85 s window, in key frequency bands of interest. Boxes indicate the interquartile range across subjects (N=12). Rightmost TFRs show the average t-statistic reflecting the Post-minus-Pre change in spectral power.

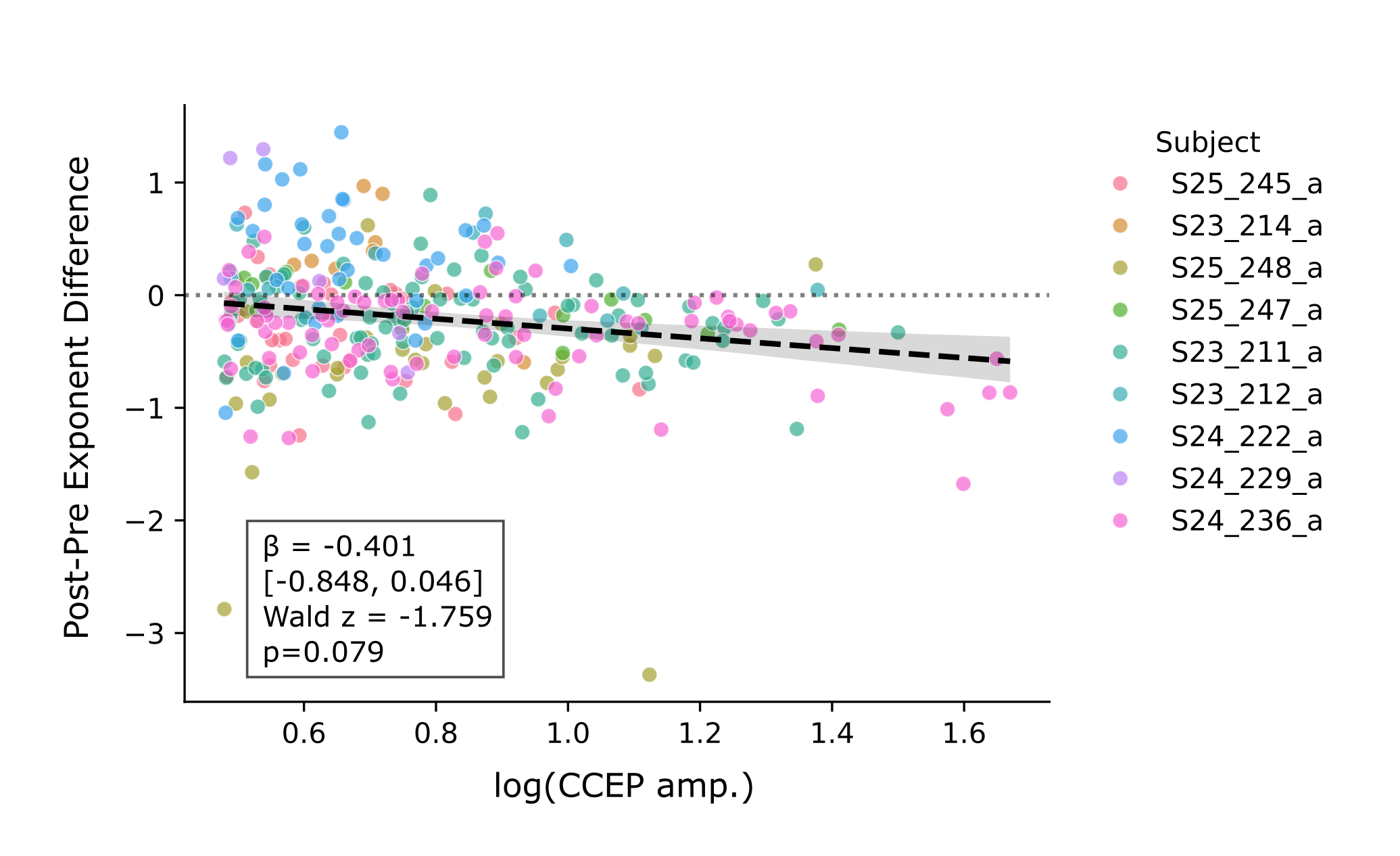

**Supplemental Figure 6. Resting-state exponent change is not significantly correlated with CCEP amplitude.** We assessed whether CCEP amplitude – a measure of effective connectivity – is correlated with iTBS-related change in resting-state exponent (main text Figure 7), focusing specifically on the third iTBS session (“iTBS3”) where significant Post-Pre iTBS exponent change was observed (Figure 7b). A *p*<0.1 trend-level effect was noted (see inset box for mixed model output and *Methods* for details). These data reflect all channels with a CCEP amplitude threshold >3, to capture variability across high and low responses.

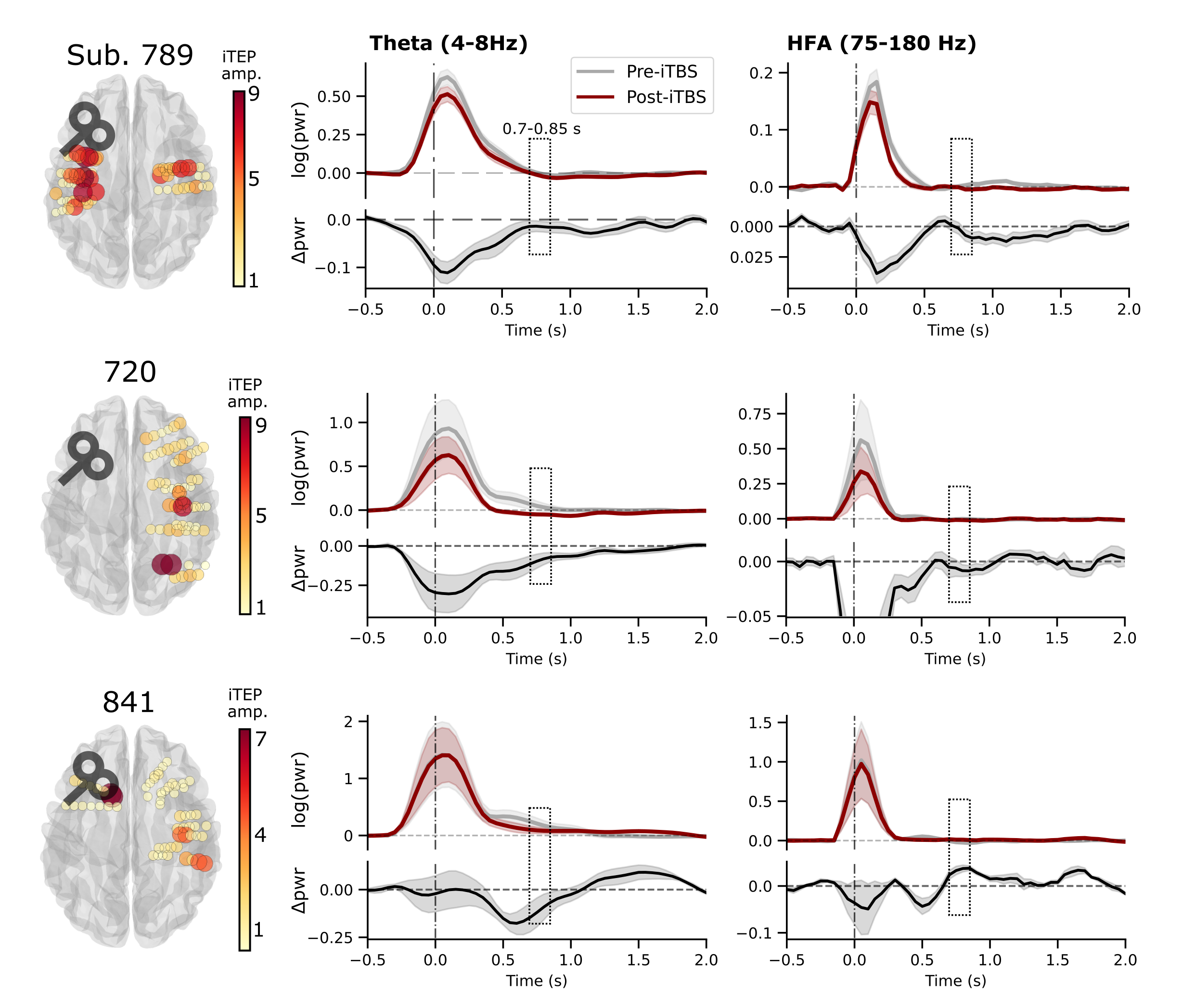

**Supplemental Figure 7. spTMS-provoked theta and HFA timecourses.** Structured identically to main text Figure 8b-c, depicted for each individual subject included in the TMS-iEEG analysis. *Left:* Brainplots display the intracranial TMS evoked potential (iTEP) amplitude, rendered on each recording channel by size and color. *Right:* spTMS-provoked power was averaged across all in-network channels for each subject (surpassing an iTEP amplitude of 3). The 0.7-0.85 s window, identified from the separate electrical stimulation experiment, is indicated. Error bars show +/- 1 SEM across channels.

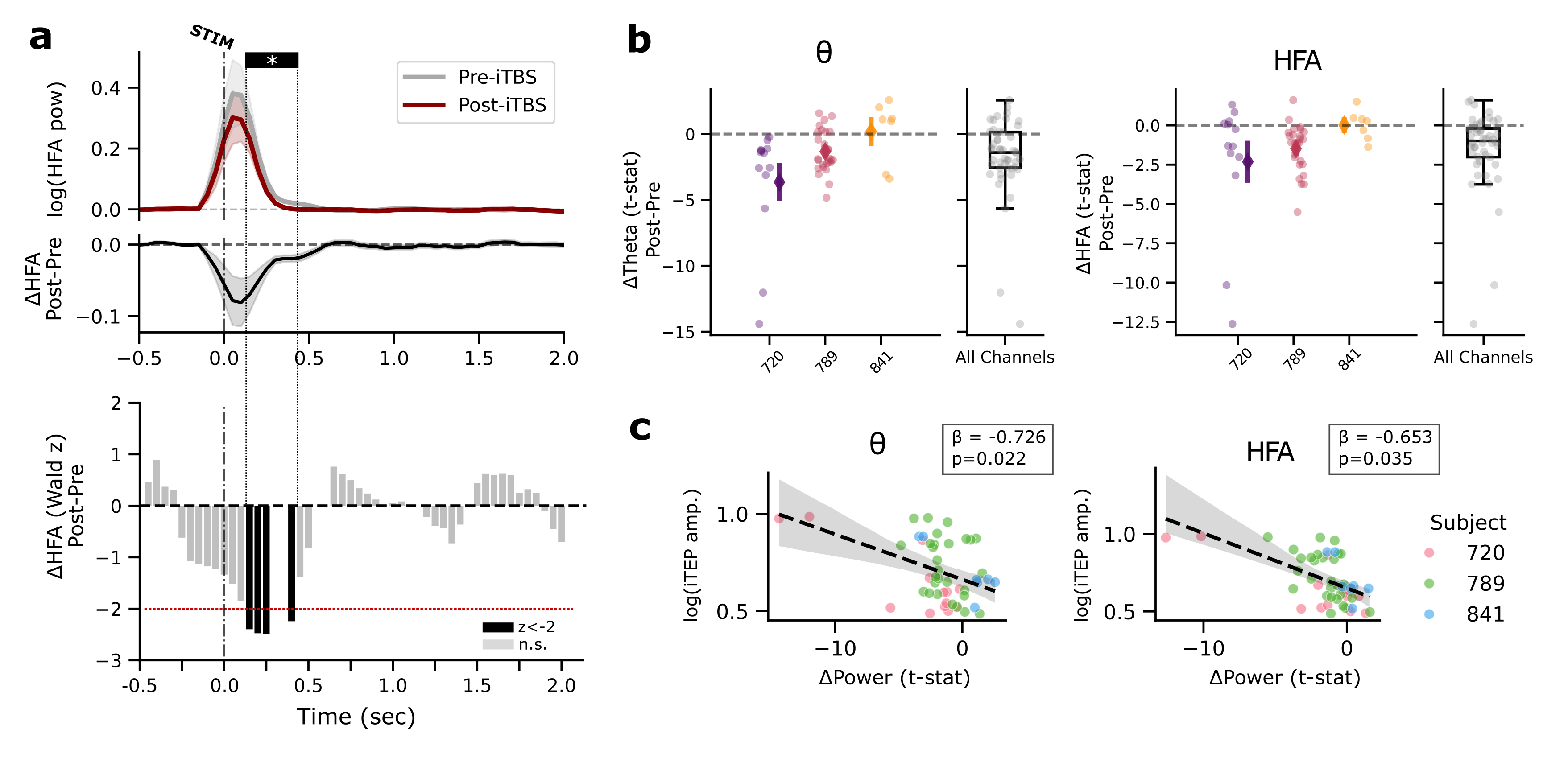

**Supplemental Figure 8. TMS-iTBS changes cortical reactivity in an early window after spTMS. (A)** To ask whether TMS-iTBS changed cortical reactivity outside the 0.7-0.85 s window derived from our electrical stimulation experiments, we re-analyzed the difference between pre and post-iTBS HFA timecourses in a manner similar to main text Figure 2b, averaging across all subjects and in-network channels. Using a mixed-effects statistical approach, we found a significant Post-Pre decrease in HFA from 0.15-0.4 s, though this result was not corrected for multiple comparisons across timepoints. An estimate of the effect at 0.3-0.35 s was omitted due to model convergence failure, likely due to the small sample size in the TMS-iTBS experiment. **(B)** As in main text Figure 8g, depicting the distribution of theta (*left*) and HFA (*right*) power values across all in-network channels for each participant, for the new 0.15-0.4 s window identified here. **(C)** As in main text Figure 8i, power *t*-statistics in the 0.15-0.4 s window were correlated with iTEP amplitude for all in-network channels across all 3 participants included in this analysis. Significant inverse relationships were detected for both theta and HFA, estimated via a mixed effects model as outlined in *Methods*.
