## Supplemental Tables for "iTBS induces a suppression-rebound pattern in human cortical excitability"

| Subject | Stim. Target Channel | Channel | CCEP amp. (absolute z) | Gray/White Stim Target | Distance (mm) | Stim Region | Channel Region |
| --- | --- | --- | --- | --- | --- | --- | --- |
| S25_245_a | LAMF8-LAMF9 | LLOF7-LLOF8 | 12.83 | Gray/White Junction | 11.15 | rostralmiddlefrontal | rostralmiddlefrontal |
| S25_248_a | LINA12-LINA13 | LHPA8-LHPA9 | 13.54 | Gray | 96.33 | superiorfrontal | middletemporal |
| S25_248_a | LINA12-LINA13 | LHPA9-LHPA10 | 13.29 | Gray | 99.15 | superiorfrontal | White Matter |
| S25_248_a | LINA12-LINA13 | LORB1-LORB2 | 23.73 | Gray | 70.32 | superiorfrontal | lateralorbitofrontal |
| S25_248_a | LINA12-LINA13 | LORB3-LORB4 | 12.41 | Gray | 58.66 | superiorfrontal | lateralorbitofrontal |
| S25_248_a | LINA12-LINA13 | LORB8-LORB9 | 12.42 | Gray | 31.60 | superiorfrontal | White Matter |
| S25_247_a | LANT15-LANT16 | LFRO12-LFRO13 | 13.08 | Gray | 25.76 | parsopercularis | parstriangularis |
| S25_247_a | LANT15-LANT16 | LFRO13-LFRO14 | 11.63 | Gray | 24.89 | parsopercularis | parstriangularis |
| S25_247_a | LANT15-LANT16 | RANT14-RANT15 | 16.28 | Gray | 110.09 | parsopercularis | parsopercularis |
| S25_247_a | LANT15-LANT16 | RANT15-RANT16 | 25.74 | Gray | 113.70 | parsopercularis | parsopercularis |
| S23_211_a | RATH9-RATH10 | RHPC9-RHPC10 | 10.18 | Gray/White Junction | 58.29 | rostralmiddlefrontal | middletemporal |
| S23_211_a | RATH9-RATH10 | RINb2-RINb3 | 11.65 | Gray/White Junction | 26.37 | rostralmiddlefrontal | insula |
| S23_211_a | RATH9-RATH10 | RINb3-RINb4 | 11.93 | Gray/White Junction | 27.54 | rostralmiddlefrontal | insula |
| S23_211_a | RATH9-RATH10 | RINb5-RINb6 | 12.11 | Gray/White Junction | 31.85 | rostralmiddlefrontal | parsopercularis |
| S23_211_a | RATH9-RATH10 | RINe2-RINe3 | 10.50 | Gray/White Junction | 28.26 | rostralmiddlefrontal | precentral |
| S23_211_a | RATH9-RATH10 | RINe5-RINe6 | 19.75 | Gray/White Junction | 22.03 | rostralmiddlefrontal | insula |
| S23_211_a | RATH9-RATH10 | RINe7-RINe8 | 15.08 | Gray/White Junction | 19.81 | rostralmiddlefrontal | insula |
| S23_211_a | RATH9-RATH10 | RPHG4-RPHG5 | 13.24 | Gray/White Junction | 67.07 | rostralmiddlefrontal | White Matter |
| S23_211_a | RATH9-RATH10 | RPHG6-RPHG7 | 16.56 | Gray/White Junction | 64.64 | rostralmiddlefrontal | bankssts |
| S23_211_a | RATH9-RATH10 | RPHG7-RPHG8 | 15.48 | Gray/White Junction | 64.38 | rostralmiddlefrontal | bankssts |
| S23_211_a | RATH9-RATH10 | RPHG11-RPHG12 | 17.27 | Gray/White Junction | 67.62 | rostralmiddlefrontal | supramarginal |
| S23_211_a | RATH9-RATH10 | RpIT3-RpIT4 | 13.12 | Gray/White Junction | 65.22 | rostralmiddlefrontal | White Matter |
| S23_211_a | RATH9-RATH10 | RpIT4-RpIT5 | 15.27 | Gray/White Junction | 68.27 | rostralmiddlefrontal | White Matter |
| S23_211_a | RATH9-RATH10 | RpIT6-RpIT7 | 22.21 | Gray/White Junction | 74.20 | rostralmiddlefrontal | inferiortemporal |
| S23_211_a | RATH9-RATH10 | RpTh1-RpTh2 | 20.83 | Gray/White Junction | 53.80 | rostralmiddlefrontal | Right-Thalamus-Proper |
| S23_211_a | RATH9-RATH10 | RpTh2-RpTh3 | 31.62 | Gray/White Junction | 52.19 | rostralmiddlefrontal | Right-Thalamus-Proper |
| S23_211_a | RATH9-RATH10 | RpTh3-RpTh4 | 12.76 | Gray/White Junction | 51.26 | rostralmiddlefrontal | Right-Thalamus-Proper |
| S23_211_a | RATH9-RATH10 | RpTh12-RpTh13 | 17.15 | Gray/White Junction | 65.08 | rostralmiddlefrontal | supramarginal |
| S23_211_a | RATH9-RATH10 | RpTh13-RpTh14 | 11.03 | Gray/White Junction | 68.17 | rostralmiddlefrontal | supramarginal |
| S23_212_a | RsDV6-RsDV7 | RPDV1-RPDV2 | 11.47 | Gray | 37.92 | rostralmiddlefrontal | superiorfrontal |
| S23_212_a | RsDV6-RsDV7 | RPDV5-RPDV6 | 12.92 | Gray | 18.83 | rostralmiddlefrontal | superiorfrontal |
| S23_212_a | RsDV6-RsDV7 | RPDV6-RPDV7 | 23.88 | Gray | 14.92 | rostralmiddlefrontal | White Matter |
| S23_212_a | RsDV6-RsDV7 | RPDV8-RPDV9 | 12.13 | Gray | 11.21 | rostralmiddlefrontal | rostralmiddlefrontal |
| S24_221_a | LANT3-LANT4 | LVOF2-LVOF3 | 13.01 | White | 47.13 | White Matter (near Putamen) | medialorbitofrontal |
| S24_221_a | LANT3-LANT4 | RVOF8-RVOF9 | 10.08 | White | 51.94 | White Matter (near Putamen) | rostralanteriorcingulate |
| S24_222_a | LACC1-LACC2 | LaIN4-LaIN5 | 10.10 | Gray | 34.29 | caudalanteriorcingulate | insula |
| S24_236_a | LOFV13-LOFV14 | LAMY12-LAMY13 | 15.55 | Gray/White Junction | 92.05 | superiorfrontal | middletemporal |
| S24_236_a | LOFV13-LOFV14 | LAMY13-LAMY14 | 16.79 | Gray/White Junction | 94.34 | superiorfrontal | middletemporal |
| S24_236_a | LOFV13-LOFV14 | LANT3-LANT4 | 11.04 | Gray/White Junction | 55.78 | superiorfrontal | Left-Thalamus-Proper |
| S24_236_a | LOFV13-LOFV14 | LHPC9-LHPC10 | 10.40 | Gray/White Junction | 91.23 | superiorfrontal | middletemporal |
| S24_236_a | LOFV13-LOFV14 | LHPC10-LHPC11 | 13.84 | Gray/White Junction | 92.57 | superiorfrontal | superiortemporal |
| S24_236_a | LOFV13-LOFV14 | LINA6-LINA7 | 17.48 | Gray/White Junction | 36.85 | superiorfrontal | White Matter |
| S24_236_a | LOFV13-LOFV14 | LINA7-LINA8 | 25.69 | Gray/White Junction | 31.56 | superiorfrontal | White Matter |
| S24_236_a | LOFV13-LOFV14 | LINA8-LINA9 | 44.62 | Gray/White Junction | 26.50 | superiorfrontal | White Matter |
| S24_236_a | LOFV13-LOFV14 | LINA9-LINA10 | 43.47 | Gray/White Junction | 21.44 | superiorfrontal | White Matter |
| S24_236_a | LOFV13-LOFV14 | LINA10-LINA11 | 37.56 | Gray/White Junction | 17.09 | superiorfrontal | White Matter |
| S24_236_a | LOFV13-LOFV14 | LINA11-LINA12 | 39.70 | Gray/White Junction | 13.55 | superiorfrontal | White Matter |
| S24_236_a | LOFV13-LOFV14 | LINA12-LINA13 | 18.00 | Gray/White Junction | 11.23 | superiorfrontal | superiorfrontal |
| S24_236_a | LOFV13-LOFV14 | LINA13-LINA14 | 18.88 | Gray/White Junction | 11.26 | superiorfrontal | superiorfrontal |
| S24_236_a | LOFV13-LOFV14 | LINA14-LINA15 | 46.74 | Gray/White Junction | 13.72 | superiorfrontal | superiorfrontal |
| S24_236_a | LOFV13-LOFV14 | LOFH1-LOFH2 | 15.39 | Gray/White Junction | 62.84 | superiorfrontal | medialorbitofrontal |
| S24_236_a | LOFV13-LOFV14 | LOFH2-LOFH3 | 21.71 | Gray/White Junction | 61.04 | superiorfrontal | medialorbitofrontal |
| S24_236_a | LOFV13-LOFV14 | LOFH3-LOFH4 | 20.66 | Gray/White Junction | 59.33 | superiorfrontal | medialorbitofrontal |
| S24_236_a | LOFV13-LOFV14 | LOFH4-LOFH5 | 17.57 | Gray/White Junction | 57.77 | superiorfrontal | White Matter |
| S24_236_a | LOFV13-LOFV14 | LOFH7-LOFH8 | 10.86 | Gray/White Junction | 54.82 | superiorfrontal | lateralorbitofrontal |
| S24_236_a | LOFV13-LOFV14 | LOFH13-LOFH14 | 12.28 | Gray/White Junction | 56.18 | superiorfrontal | parstriangularis |
| S24_236_a | LOFV13-LOFV14 | RINA14-RINA15 | 23.89 | Gray/White Junction | 32.05 | superiorfrontal | superiorfrontal |
| S24_236_a | LOFV13-LOFV14 | ROFV13-ROFV14 | 12.78 | Gray/White Junction | 26.72 | superiorfrontal | superiorfrontal |
| S24_236_a | LOFV13-LOFV14 | ROFV15-ROFV16 | 23.78 | Gray/White Junction | 26.43 | superiorfrontal | superiorfrontal |
| S25_120c_b | RLIP2-RLIP3 | RCRO5-RCRO6 | 10.92 | Gray | 9.57 | superiorparietal | precuneus |
| S25_120c_b | RLIP2-RLIP3 | RCRO6-RCRO7 | 15.95 | Gray | 6.40 | superiorparietal | precuneus |
| S25_120c_b | RLIP2-RLIP3 | RPPR5-RPPR6 | 12.56 | Gray | 14.34 | superiorparietal | precuneus |
| S25_120c_b | RLIP2-RLIP3 | RPPR6-RPPR7 | 13.55 | Gray | 12.57 | superiorparietal | precuneus |
| S24_220_a | LMCC2-LMCC3 | RMCC4-RMCC5 | 12.11 | Gray | 21.87 | superiorfrontal | caudalanteriorcingulate |
| S26_263_b | RAMY1-RAMY2 | RANT1-RANT2 | 11.86 | Gray | 27.06 | Right-Amygdala | Right-Thalamus-Proper |
| S26_263_b | RAMY1-RAMY2 | RLMD10-RLMD11 | 11.16 | Gray | 28.07 | Right-Amygdala | insula |
| S26_263_b | RAMY1-RAMY2 | RPLV2-RPLV3 | 11.41 | Gray | 37.85 | Right-Amygdala | Right-Thalamus-Proper |
| S26_263_b | RAMY1-RAMY2 | RPLV3-RPLV4 | 12.10 | Gray | 38.45 | Right-Amygdala | Right-Thalamus-Proper |
| S26_263_b | RAMY1-RAMY2 | RPLV4-RPLV5 | 14.23 | Gray | 39.53 | Right-Amygdala | White Matter |
| S26_263_b | RAMY1-RAMY2 | RPLV5-RPLV6 | 13.76 | Gray | 41.41 | Right-Amygdala | White Matter |
| S26_264_a | RINP15-RINP16 | RPLV2-RPLV3 | 11.34 | Gray | 45.44 | superiorparietal | Right-Thalamus-Proper |

**Table 1. Recording channels included in primary analysis (CCEP threshold >10).** Columns include subject ID (letter code refers to multiple stimulation sites within a single subject), stimulation target channel ID, recording channel ID, CCEP amplitude (z-score units), location of stimulation site relative to gray matter, white matter, or gray/white boundary, distance in mm (MNI space) between the recording and stimulation channel, stimulation region (Desikan-Killiany atlas labels), and channel region.

| **Figure** | **# Subjects** | **Mixed Effects Model Specification** |
| --- | --- | --- |
| 2b-d | N=12; effects pooled across all iTBS sessions | 1. Intercept-only model for each 200ms time window, FDR corrected: channel_t_stat ~ block + (1 \| subject) 2. For comparisons within modulation window: channel_t_stat ~ block + network_category + active_vs_sham + (1 \| subject) |
| 3a-b | N=12 (>10 threshold), N=15 (>5 threshold); effects pooled across all iTBS sessions | No new model tested |
| 3d | N=17 (>3 threshold); effects pooled across all iTBS sessions | No random intercept as data already centered per subject, no block category as data averaged across blocks: channel t-stat ~ ptp_amplitude + distance + (0 + ptp_amplitude + distance \| subject) |
| 4a | N=12; effects pooled across all iTBS sessions | Intercept-only model per frequency band, Bonferroni corrected: channel_t_stat ~ block + (1 \| subject) |
| 4b-c | N=12 (iTBS1), N=9 (iTBS2), N=9 (iTBS3) | 1. For each individual iTBS block, Intercept-only model: channel_t_stat ~ 1 + (1 \| subject) 2. Testing for linear progression over blocks: Δpower ~ block_number + (1 \| subject) |
| 5a-c | N=12 (iTBS1), N=9 (iTBS2), N=9 (iTBS3) | 1. For each individual iTBS block or sham block, Intercept-only model: channel_t_stat ~ 1 + (1 \| subject) 2. For average effect across all blocks: channel_t_stat ~ block + (1 \| subject) |
| 6b | N=12 (iTBS1), N=9 (iTBS2), N=9 (iTBS3) | 1. Intercept-only model per block: Δpower ~ 1 + (1 \| subject) 2. Across blocks: Δpower ~ block + (1 \| subject) 3. For linear progression across blocks: Δpower ~ block_number + (1 \| subject) |
| 6c | N=8 (those who completed all three iTBS sessions with limited timing deviations) | Intercept-only model for testing deviation from baseline, per block: power ~ 1 + (1 \| subject) |
| 7 | N=9 unique subjects total (those with substantial resting-state time available); N=6 (iTBS1), N=6 (iTBS2), N=7 (iTBS3) | 1. Effect across blocks and effect of network category: Δexponent ~ block + network_category + (1 \| subject) 2. Per individual block: paired *t*-test due to model convergence issues at low Ns. |
| 8f, 8i | N=3 (patients who underwent TMS-iTBS) | 1. Average effect: channel_t_stat ~ 1 + (1 \| subject) 2. Correlation: channel_t_stat ~ ptp_amplitude + (0 + ptp_amplitude \| subject) |

**Table 2. Number of subjects included in each analysis** **and mixed model specification for key tests.**
